# Infoveillance study on the dynamic associations between CDC social media contents and epidemic measures during COVID-19

**DOI:** 10.1101/2023.06.26.23291921

**Authors:** Shuhua Yin, Shi Chen, Yaorong Ge

## Abstract

**Background:** Health agencies have been widely adopting social media to disseminate important information, educate the public on emerging health issues, and understand public opinions. The Centers for Disease Control and Prevention (CDC) has been one of the leading agencies that utilizes social media platforms during the COVID-19 pandemic to communicate with the public and mitigate the disease in the United States. It is crucial to understand the relationships between CDC’s social media communication and the actual epidemic metrics to improve public health agencies’ communication strategies during health emergencies.

**Objective:** The aim of this study was to identify key topics in tweets posted by CDC during the pandemic, to investigate the temporal dynamics between these key topics and the actual COVID-19 epidemic measures, and to make recommendations for CDC’s digital health communication strategies for future health emergencies.

**Methods:** Two types of data were collected: 1) a total of 17,524 COVID-19-related English tweets posted by the CDC between December 7, 2019 and January 15, 2022; 2) COVID-19 epidemic measures in the U.S. from the public GitHub repository of Johns Hopkins University from January 2020 to July 2022. Latent Dirichlet allocation (LDA) topic modeling was applied to identify key topics from all COVID-19-related tweets posted by CDC, and the final topics were determined by domain experts. Various multivariate time series analysis techniques were applied between each of the identified key topics and actual COVID-19 epidemic measures to quantify the dynamic associations between these two types of time series data.

**Results:** Four major topics from CDC’s COVID-19 tweets were identified: 1) information on prevention of health outcomes of COVID-19; 2) pediatric intervention and family safety; 3) updates of the epidemic situation of COVID-19; 4) research and community engagement to curb COVID-19. Multivariate analyses showed that there were significant variabilities of progression between CDC’s topics and the actual COVID-19 epidemic measures. Some CDC’s topics showed substantial associations with the COVID-19 measures over different time spans throughout the pandemic, expressing similar temporal dynamics between these two types of time series data.

**Conclusions:** Our study is the first to comprehensively investigate the dynamic associations between topics discussed by CDC on Twitter and the COVID-19 epidemic measures in the U.S. We identified four major topic themes via topic modeling and explored how each of these topics was associated with each major epidemic measure by performing various multivariate time series analyses. We recommend that it is critical for public health agencies, such as CDC, to disseminate and update timely and accurate information to the public and align major topics with the key epidemic measures over time. We suggest that social media can help public health agencies to inform the public on health emergencies and to mitigate them effectively.

## Introduction

The COVID-19 pandemic had caused more than 760 million cases and 6.8 million deaths globally as of April 2023 [1]. Therefore, it is crucial for public health agencies such as the U.S. Center for Disease Control and Prevention (CDC) to quickly and effectively disseminate up-to-date and reliable health information to the public to curb the pandemic. Over the past years, social media has been widely used by various public health agencies to make announcements, disseminate information, and deliver guidelines of effective interventions to the public. CDC is among the early adopters of social media to engage with the public, increase health literacy in the society, and promote healthy behaviors [2]. Moreover, CDC’s social media team has developed the Health Communicator’s Social Media Toolkit to efficiently utilize social media platforms, map health strategies, listen to health concerns from the public, and deliver evidence-based, credible, and timely health communications in multiple formats such as in texts, images, and videos. CDC’s digital health communication efforts have been especially established on various social media platforms such as Twitter, Facebook, and Instagram.

Building successful interactions with the public relies on people understanding the content and raising awareness of it. CDC has been heavily engaging in social media presence [3]. For example, during the COVID-19 pandemic since 2019, it has been responsive and proactive on Twitter to continuously tweet about reliable health-related messages and quickly diffuse public engagement by responding to user comments, retweeting credible sources, and monitoring online conversations in real time. Hence, it is meaningful to recognize the COVID-19 pandemic information disseminated by CDC on social media, characterize various contents and topics, and evaluate posting patterns with regard to the actual epidemic dynamics. Monitoring the content, topics, and trends will help identify current issues or interests and levels of interventions. It is critical to evaluate the associations between various COVID-19 content topics tweeted by CDC and the actual COVID-19 epidemic measures (e.g. cases, deaths, testing and vaccination records). Knowing the underlying associations between CDC’s digital health communication contents on social media, and the actual COVID-19 epidemics will help understand and evaluate CDC’s tweeting patterns with the changes of the epidemic, and further recommend more effective social media communication strategies for public health agencies accordingly.

Infodemiology and infoveillance studies target on solving health problems, analyzing insights and dynamics, and predicting patterns and trends of diseases using online data. Infodemiology, which is the conjunction of “information” and “epidemiology”, defined by Gunter Eysenbach, is the field of distribution and determinants of information of a population through Internet or other electronic media [4]. Infoveillance takes surveillance as the primary aim and generates automated analysis from massive online data. It employs innovative methods and approaches to mine and analyze unstructured online text information such as analyzing patterns, trends, make predictions for future events such as potential outbreak, and help address current underlying issues of public health. Instead of traditional epidemiological surveillance systems, which include cohort studies, disease registries, population surveys, and healthcare records, etc., infoveillance studies discover wide range of health topics, monitor health issues including outbreaks or pandemics, and forecast epidemiological trends in real time. Large amount of anonymous online data can be obtained in a much timely manner than the traditional surveillance systems, and this will help researchers and public health officials to prepare for and tackle public health emergencies and issue more efficiently and effectively.

Social media platforms have been making its impact on community education of COVID-19 and delivering various health information about the disease. Many studies have also incorporated the concept of infoveillance by analyzing unstructured textual data obtained from social media platforms and gaining insights from the results. Liu et al collected and analyzed media reports and news articles on COVID-19 to derive topics and useful information [5]. They aimed to investigate the relationship between media reports and the COVID-19 outbreak, and the patterns of health communication on coronavirus through mass media to the general audience. They obtained the media reports and articles related to the pandemic and studied prevalent topics.

There had been prevalent public discussions of attitudes and perspectives on mask-wearing on social media. Therefore, it is important for public health agencies to disseminate supporting evidences on the benefits of masking to mitigate the spread of COVID-19. Al-Ramahi et al studied the topics associated with the public discourse against wearing masks in the United States on Twitter [6]. They identified and categorized different topics in their models. These studies all aimed to apply infoveillance to investigate potential impacts of diseases, health behaviors or interventions on target populations, communities, and the society. However, mass and social media are also prone to the spreading of misinformation and conspiracy theories, especially from unreliable sources [7]. Hence, the sources of information obtained from social media is crucial as social media could contain misinformation and potentially create bias and mislead public perceptions and emotions. Nevertheless, official public health agencies’ accounts are usually sources of unbiased and reliable health information. Although there have been several studies that collectively explored the topics discussed by the general public on social media during the pandemic, no study attempts have been made to identify various topics from health agencies such as CDC so far.

Furthermore, information discrepancies and delays could occur between topics posted by health agencies and real-time epidemic trends. Such discrepancies could potentially cause confusions to the public on interventions for health emergencies. Therefore, quantifying their relationships is important to reduce knowledge gaps. Chen et al studied dynamics between Zika epidemic in 2016 and CDC’s response on Twitter [8]. They quantified the association between the two types of data through multivariate time series analysis and information theory measurements. The study discovered CDC’s varying degrees of efforts in disseminating health-related information to the public during different phases of the Zika pandemic in 2016. However, no study so far has investigated such dynamic associations, more specifically, CDC’s COVID-19 content topic tweeting patterns and the actual COVID-19 epidemic metrics.

Understanding the dynamic association between social media content topics and epidemic metrics will help health agencies to identify driving factors between the two and disseminating helpful knowledge to the public accordingly. In this study, we aim to discover the underlying COVID-related topics posted by CDC during different phases of the COVID-19 pandemic. We also aim to further quantify and evaluate the dynamic associations between content topics of the pandemic and multiple COVID-19 epidemic metrics. This study will significantly increase our insights about the efficiency of CDC’s health communications during the pandemic and make recommendations for CDC’s social media communication strategies with the public in the future.

## Methods

### Data Acquisition and Preprocessing

Using the Twitter academic API and search query (see search query in Appendix), we retrieved a total of 17,524 English tweets posted by seven official CDC-affiliated Twitter accounts to January 15, 2022. These accounts are: @CDCgov: CDC’s official Twitter source for daily credible health and safety updates from Centers for Disease Control & Prevention; @CDCEmergency: CDC Emergency, which tweets ways to for public health preparedness during emergency responses; @CDCDirector: account of the CDC director of the time; @CDCGlobal: CDC Global health, which tweets about how CDC strives to contribute to saving lives, reducing disease, and improving global health around the world; @CDCtravel: CDC Travel health, which voices to help travelers and their clinicians prevent illness and injury during international travel; @DrKhabbazCDC: past Twitter account from former director of CDC emerging infections (NCEZID), Dr. Rima Khabbaz. NCEZID works to protect people from emerging and zoonotic infectious diseases, from anthrax to Zika; @CDCMMWR: MMWR (Morbidity and Mortality Weekly Reports), which is CDC’s primary vehicle for scientific publication of timely, authoritative, and useful public health information and recommendations. We consolidated these tweets into a data frame of daily tweet counts of each of the seven CDC-affiliated accounts during the time period. The associated metadata include tweet posting dates, textual data of the tweets, tweet account ID, account types (organization or individual), public engagement metrics (e.g., number of retweets, replies, likes, and quotes), referenced tweet type (retweeted, replied to, and quoted), and referenced tweet IDs.

We also acquired the COVID-19 epidemic metric data in the U.S. from Johns Hopkins University – Center for Systems Science and Engineering (CSSE) public GitHub repository ([13]–[15]). Four sets of important COVID-19 time series data were retrieved, including daily cumulative confirmed cases, deaths, testing, and vaccination. The data were all at U.S. nation-level. The original four sets of COVID-19 time series data consist of dates and their cumulative targeted measurements. Case series includes daily cumulative number of confirmed COVID-19 reported cases, and it has 751 records, ranging from January 22, 2020 to February 10, 2022.

Death series reports daily cumulative number of confirmed COVID-19 death cases, and it has 908 records, ranging from January 22, 2020 to July 17, 2022. Testing data reports daily cumulative number of completed PCR tests or other approved nucleic acid amplification test (NAAT), and it has 760 records, ranging from January 13, 2020 to February 10, 2022. Vaccination data includes daily cumulative number of people who received a complete primary series of vaccine doses, from the CDC Vaccina Tracker; vaccination series has 428 records, ranging from December 10, 2020 to February 10, 2022.

For consistency in analysis, all CDC tweets time series and U.S. COVID-19 variable time series were standardized to the same time span in this study, ranging from the start-date of reported cases data, January 22, 2020 (the start-date of reported cases data), to the end-date of CDC tweet collection on January 15, 2022, with total of 725 records for each data. Since vaccination data were not available until late 2020, missing values in the vaccination series were filled with 0s. In summary, we have four time series from four different COVID-19 U.S. epidemic metrics, and another social media time series of tweets from all seven CDC-associated Twitter accounts.

### Natural Language Processing (NLP)

In order to identify major topics in CDC’s COVID-19 tweets, we performed various natural language preprocessing (NLP) analyses. NLP, especially topic modeling, provides granular characterization of textual inputs such as CDC’s COVID-19 communications.

Regular expressions were first applied to process tweet texts by removing @mentions, hashtags, special characters, emails, punctuations, URLs, and hyperlinks. Tokenization was performed to break down sentences into individual tokens, which can be individual words or punctuations.

For example, the sentence “As COVID19 continues to spread, we must remain vigilant.” becomes tokens of “As”, “COVID19”, “continues”, “to”, “spread”, “,”, “we”, “must”, “remain”, “vigilant” after tokenization. Next, lemmatization, a structural transformation where each word or token was turned to its base or dictionary form of their morphological information, was performed. For example, for words “studies” and “studying”, their base form, or lemma, was the same “study”. In addition to stop words removal via Python NLTK library, we also created our own list of stop words and removed them from the texts (see stopwords list in Appendix). With help from domain experts, we excluded top words that did not contribute to topic mapping.

N-grams, phrases with *n* words, were developed with a threshold value of 1 to form phrases from tweets. Phrase-level *n*-grams were applied here because phrases offer more semantic information than individual words [16]. Higher threshold value resulted in fewer phrases to be formed. The texts were mapped into a dictionary of word representations, which was a list of unique words, and was then used to create bag-of-words presentations of the texts. A TF-IDF (term frequency-inverse document frequency) model was implemented to evaluate the importance and relevancy of the words to a document. It was calculated by multiplying term frequency, which was the relative frequency a word within a document, with the inverse document frequency, which measures how common or rare a word is across a corpus. Higher TF-IDF values indicates that the word is more relevant to the document it is in ([17], [18]). Words that were missing and lower than the threshold value 0.005 from the TF-IDF model were excluded. Figure 1 shows the process of data collection and pre-processing, and Figure 2 shows the steps of subsequent NLP and statistical analyses.

**Table 1.** Data collection and preprocessing.

| Data Collection |  | Data Preprocessing |
| --- | --- | --- |
| CDC Tweets | ➤ Twitter API using search query<br>➤ 17,524 English tweets by Jan 15, 2022 | ➤ Remove @mentions, hashtags, special characters, emails, punctuations, URLs, and hyperlinks<br>➤ Tokenization: break down sentences into individual tokens<br>➤ Lemmatization: each word or token was turned to its base or dictionary form<br>➤ Remove list of stop words created by research experts<br>➤ N-grams: form phrases from the tweets<br>➤ Modify the date range: Jan 22, 2020 (the start-date of reported cases data) - Jan 15, 2022 (to the end-date of CDC tweets) |
| COVID-19 Epidemic Metrics | ➤ Public GitHub repository of CSSE at Johns Hopkins University:<br>1. Confirmed case counts: 751 records, Jan 22, 2020 - Feb 10, 2022<br>2. Death counts: 908 records, Jan 22, 2020 - Jul 17, 2022<br>3. Completed COVID-19 tests: 760 records Jan 13, 2020 - Feb 10, 2022<br>4. Complete vaccinations: 428 records, Dec 10, 2020 - Feb 10, 2022 | ➤ Standardize metrics time series to be the same as that of CDC tweets<br>➤ Fill missing values in the vaccination data with 0s<br>➤ 725 records for each of the four metrics series<br>➤ Turn cumulative to daily records |

**Table 2.**
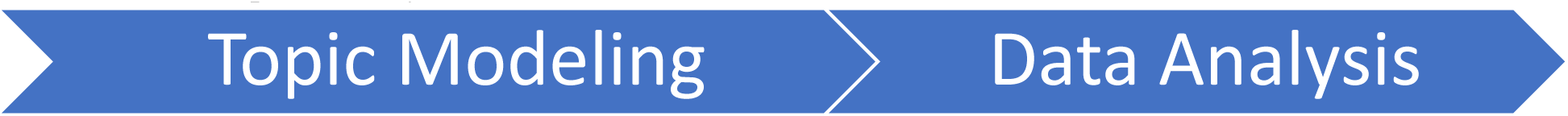
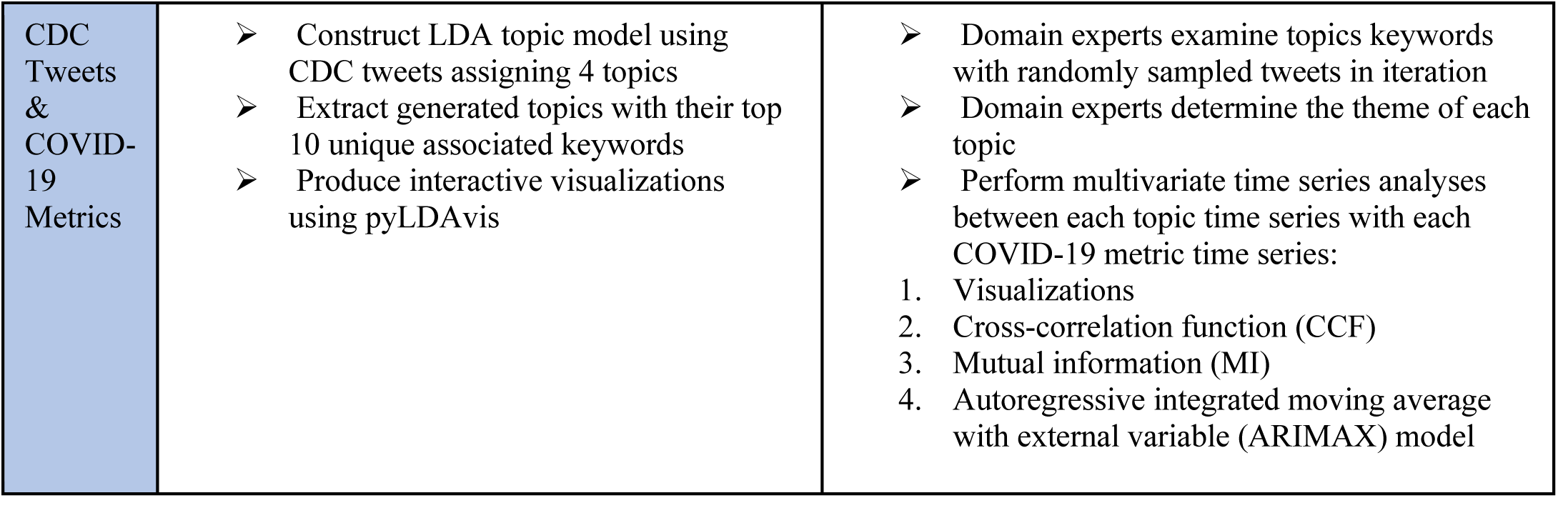
Subsequent analyses.

|  |  |  |
| --- | --- | --- |
| CDC Tweets & COVID-19 Metrics | ➤ Construct LDA topic model using CDC tweets assigning 4 topics<br>➤ Extract generated topics with their top 10 unique associated keywords<br>➤ Produce interactive visualizations using pyLDAvis | ➤ Domain experts examine topics keywords with randomly sampled tweets in iteration<br>➤ Domain experts determine the theme of each topic<br>➤ Perform multivariate time series analyses between each topic time series with each COVID-19 metric time series:<br>1. Visualizations<br>2. Cross-correlation function (CCF)<br>3. Mutual information (MI)<br>4. Autoregressive integrated moving average with external variable (ARIMAX) model |

**Figure 1.**
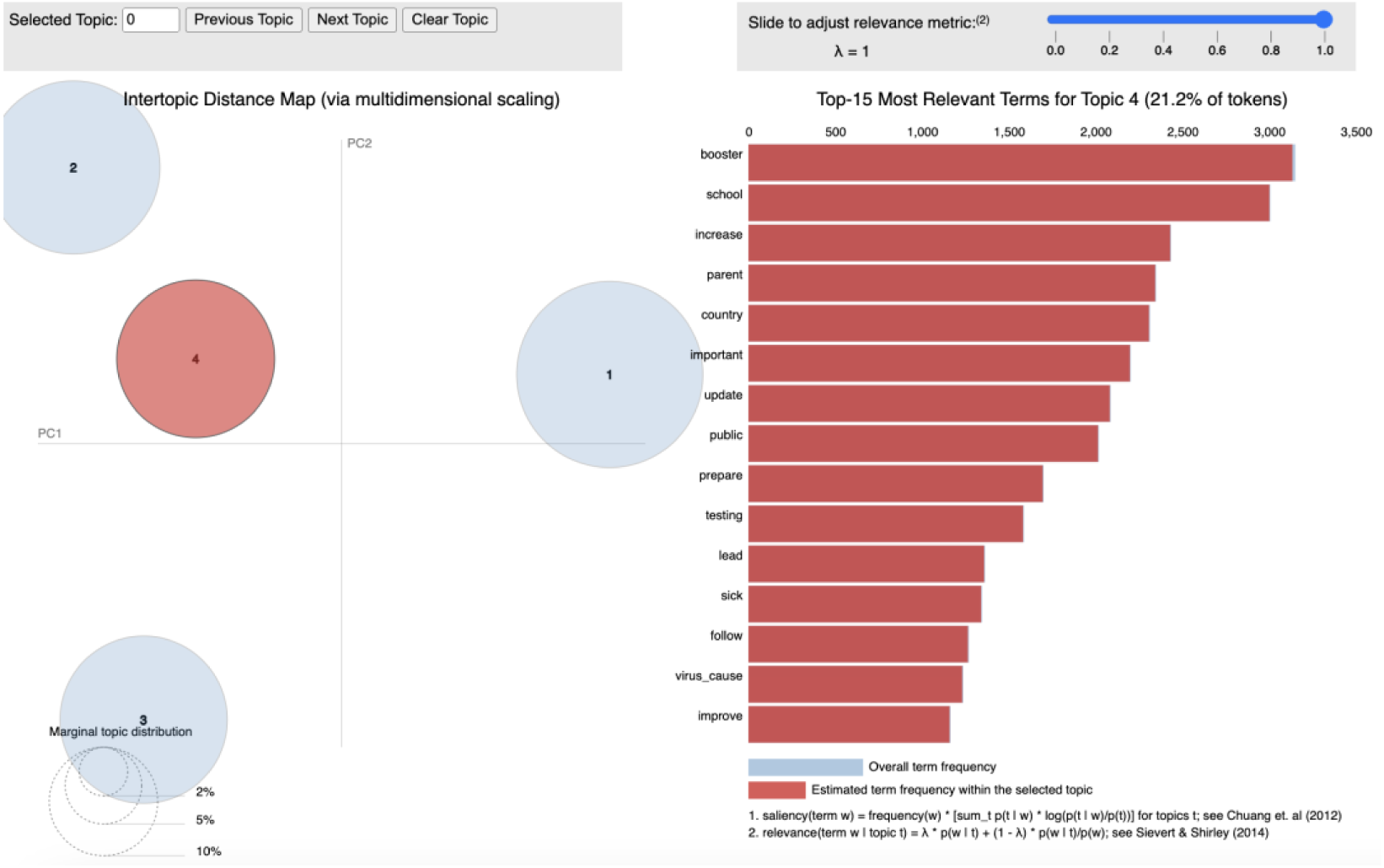
Interactive mapping of topic 4 generated by LDA

**Figure 2.**
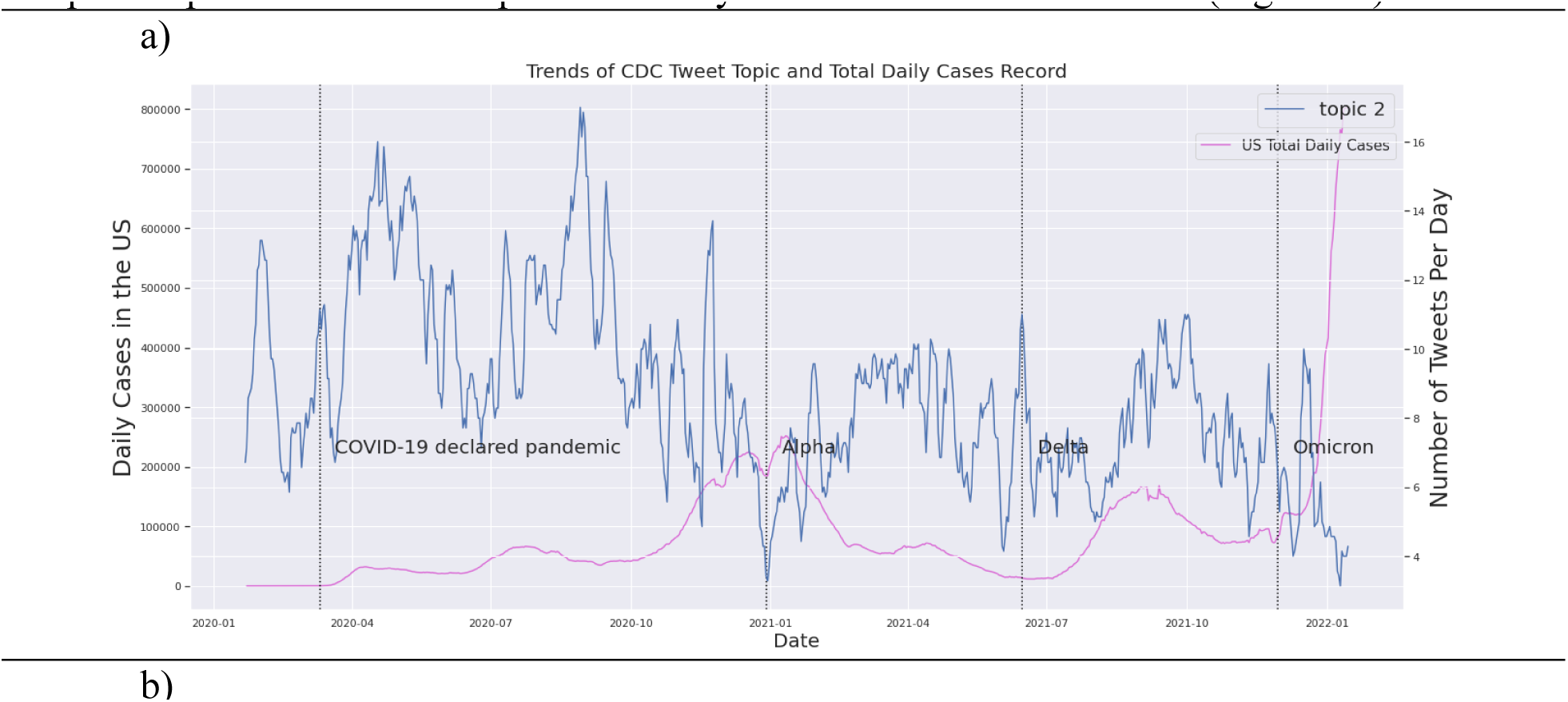
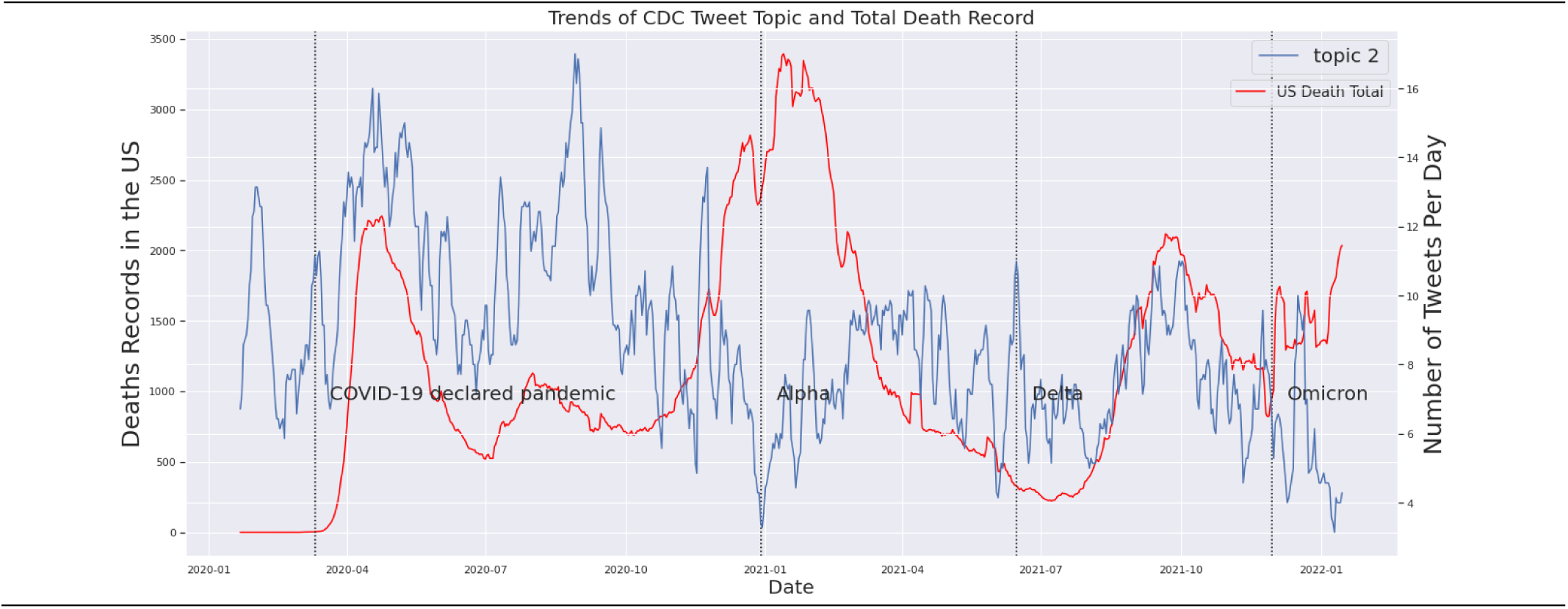
Time series of topic 2 against two COVID-19 metrics: a) confirm case counts, b) death counts

### Topic Modeling with Latent Dirichlet Allocation (LDA)

To identify more specific topics from all the COVID-19 tweets posted by CDC, we performed topic modeling via latent Dirichlet allocation (LDA). LDA automatically generates non-overlapping clusters of distributions of words that represent different topics based on probabilistic distributions. LDA was developed to find latent, or hidden topics from a collection of unstructured documents, or corpus with text data. Topic models are probabilistic models that performs on three levels of documents: a word, a document, and a corpus. Details of LDA and topic models are provided in supplementary information (SI). We investigated and compared across 3 to 8 potential topics, and determined the optimal number of topics based on both topic model evaluation and domain expert interpretations of the identified topic clusters.

Model perplexity and topic coherence scores were calculated as performance metrics of LDA. Perplexity is a decreasing “held-out log-likelihood” function that assesses LDA performance using a set of training documents. The trained LDA model is then used to test documents (held-out set). The perplexity of a probability model *q* on how well it predicts a set of samples x*_1_*, x*_2_*, …, x*_N_* drawn from an unknown probability distribution *p,* is defined as:

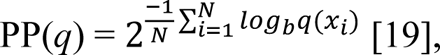

An ideal *q* should have high probabilities q(x*_i_*) for the new data. Perplexity decreases as the likelihood of the words in new data increases. Therefore, lower perplexity indicates better predictability of an LDA model.

Topic coherence assesses the quality of the topics, measured as the understandability and semantic similarities between high scoring words (i.e., the words that have high probability of occurring within a particular topic) in topics generated by LDA [20]. We used UMass coherence score [21], which accounts for the order of a word appears among the top words in a topic. It is defined as:

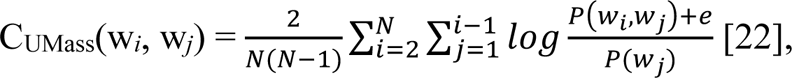

where *N* = the number of top words of a topic of a sliding window, *P*(w*_i_*) is the probability of the *i*-th word w appearing in the sliding window that moves over a corpus to form documents, *P*(w*_i_*, w*_j_*) is the probability of words w*_i_* and w*_j_* appear together in the sliding window. According to the study from UMass, coherence decreases initially and becomes stationary as the number of topics increases [20].

Human domain experts are involved to further interpret the generated topics. Representations of all topics were presented in word-probability pairs for the most relevant words generated by the topics. Interactive visualizations were produced using pyLDAvis package in Python 3.7 to examine the topics generated by LDA and their respective associated keywords. A data frame of all dominant, key topics was created. The original unprocessed full texts of the CDC tweets, IDs, and posting date were combined into a data frame along with their corresponding key topic number label and topic keywords. In addition, daily percentage of each topic from LDA were also calculated for further time series analysis. For instance, vaccine/vaccination is an identified key topic in CDC’s COVID-19 tweets, so the percentage of vaccine-related tweets in each day was calculated for the entire study period to construct the vaccine/vaccination specific topic time series. Since LDA is technically an unsupervised clustering method, we further labeled and interpreted the topics using domain knowledge. Four sets, each containing 20 tweet samples from each identified topic were randomly selected. Domain experts then examined and discussed the sampled tweets from each topic and labeled the topics (e.g., vaccine/vaccination). The final agreement on the interpretation of LDA-generated topics were reached after multiple iterations and discussions of the above process.

### Multivariate Time Series Analyses between Identified CDC Tweet Topics and COVID-19 Epidemic Metrics

#### Data preparation

Key topic time series data were derived from the previous NLP and LDA processes. We constructed a multivariate data frame with posting dates and number of tweets for each key topic at daily resolution. Since LDA identified 4 key topics, a total of four CDC key topic time series were developed. The four COVID-19 epidemic metric time series data frame contained dates and daily cumulative reported cases, cumulative confirmed deaths, cumulative number of completed PCR tests or other approved nucleic acid amplification test (NAAT), and cumulative number of people who received a complete primary series of vaccines. These four sets of COVID-19 epidemic time series were then converted from cumulative to daily measures via first order differencing for the purpose of our analysis. Multivariate time series analyses were implemented to investigate the associations between time series of key CDC tweet topics and U.S. COVID-19 epidemic metrics.

#### Visualizations

Both types of time series, CDC key topics and COVID-19 metrics were displayed in the same plot for visualization and inspection on double y-axes, with left y-axis displaying the daily COVID-19 metric, and right y-axis displaying daily CDC tweet topic counts. In addition, each plot was further sectioned based on COVID-19 phases with different dominant variant: the original, Alpha, Delta, and Omicron, with their corresponding starting dates: March 11, 2020; December 29, 2020; June 15, 2021; and November 30, 2021. This helps further observe and identify dynamic changes over time.

#### Cross-Correlation Function (CCF)

Between two time series (also known as signals x and y), cross-correlation functions (CCF) [24] quantify their levels of similarities (i.e., how similar the two series at different times) and associations (i.e., how values in one series can provide information about the other series) and examine when they occur [23]. CCF takes the sum of product for each of x and y data points at time lag *l*, defined as below:

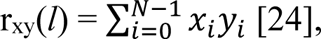

where *N* is the number of observations in each time series; x*_i_* and y*_i_* are the observations at *i*-th time step in each of their time series. CCF ranges from -1 to 1, and the larger the absolute values of CCF, the higher association the two time series share at a given time lag *l* [25]. In this study, each of the four CDC tweet topic time series was compared with each of the four COVID-19 epidemic metrics time series to calculate their respective CCFs. All values of CCFs were calculated with a maximum lag of 30 days, as we assumed that the real-world disease outcome could not influence online discussions for a month, and vice versa.

#### Mutual Information (MI)

Mutual information (MI) was calculated by computing the entropy of the empirical probability distribution to further quantify the association between each of the four key CDC tweet topics and each of the four U.S. COVID-19 epidemic metrics. MI measures the amount of mutual dependence, or average dependency between two random variables X, and Y. It is defined as the following equation:

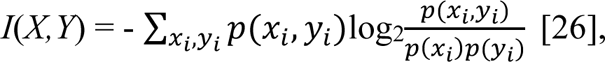

where x_i_, y_i_ are the i-th element of the variables X and Y, respectively. When applied to time series data, X and Y are two individual time series and x_i_ and y_i_ are their respective observations at *i*-th time step. Note that MI is a single value instead of a function over lag *l* as in CCF. Larger MI value indicates higher shared mutual dependency between the two time series.

#### Autoregressive Integrated Moving Average with External Variable (ARIMAX)

Neither CCF nor MI differentiate dependent and independent variables, i.e., the formula was symmetric with regard to X and Y variables. We further evaluated whether the CDC tweeting topics as dependent variables corresponded to the changes of real-world COVID-19 epidemic metrics. An Autoregressive Integrated Moving Average with External Variable (ARIMAX) model was constructed to fit each of the four CDC topics with each of the four COVID-19 epidemic metrics during the entire study period. Univariate Autoregressive integrated moving average (ARIMA) model fits and forecasts time series data with an integration of autoregressive (AR) component and a moving average (MA) component with their respective orders/lags (see supplementary materials for detailed information about AR model). The ARIMA model consists of both AR(*p*) and MA(*q*) as well as order *d* differencing term, resulting in the following ARIMA (*p*, *d*, *q*) model:

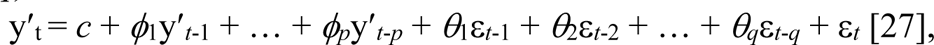

or in backward shift form:

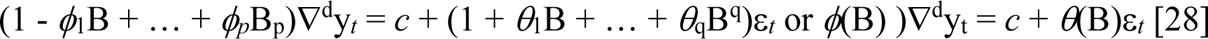

(see supplementary materials for details on the parameters).

The ARIMAX model further extends ARIMA to multivariate time series by incorporating at least one exogenous independent variable x*_t_*. The ARIMAX (*p*, *d*, *q*) is specified as:

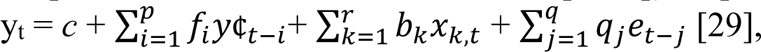

or in backward shift operator form:

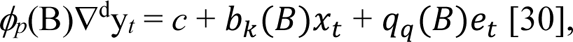

Where 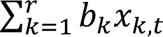 contributes to the exogeneous independent variable that could potentially influence the dependent variable y_t_.

In this study, ARIMAX was developed to evaluate how real-world epidemic metrics, modeled as exogeneous variables, impact CDC tweet topic dynamics as dependent variable. Each one of the four CDC tweet topics was modeled as dependent variable (y_t_) and each of the four COVID-19 epidemic measures was independent exogeneous variable (x_t_). The optimal ARIMA and ARIMAX model parameter set (*p, d, q*) was determined by the R ARIMA model package.

In addition to reporting the values of ARIMAX model parameters *p*, *d*, and *q*, dAIC, RMSE, and MAE were computed to compare ARIMAX performances. The optimal model was the one with the lowest AIC score. dAIC (difference in Akaike information criterion) was computed in between two models (see supplementary materials for detailed information on AIC). ARIMA model of a single topic time series and ARIMAX model of that topic time series with an exogeneous variable. Negative dAIC values indicated that ARIMAX model has improvement in model performance over the ARIMA counterpart that did not include an exogenous variable.

Commonly used root-mean-squared error (RMSE) and mean absolute error (MAE) are adopted as performance metrics. They are defined as:

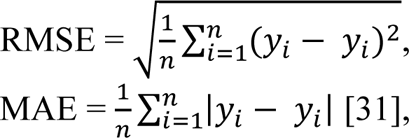

where *n* = the number of data points in a sample y (y*_i_*, where *i* = 1, 2, …, *n*). RMSE and MAE are Euclidean distance and Manhattan distance in high-dimensional space, respectively.

## Results

### Topic Modeling and Content Results

A total of 17,524 English tweets posted by CDC were retrieved and analyzed. LDA topic model generated 4 key topics based on the reference evaluation scores (i.e., perplexity and coherence score). These topics were then examined and categorized to themes by domain experts. The themes of the topics and their top 10 unique associated keywords are presented in the table below:

**Table 3.**
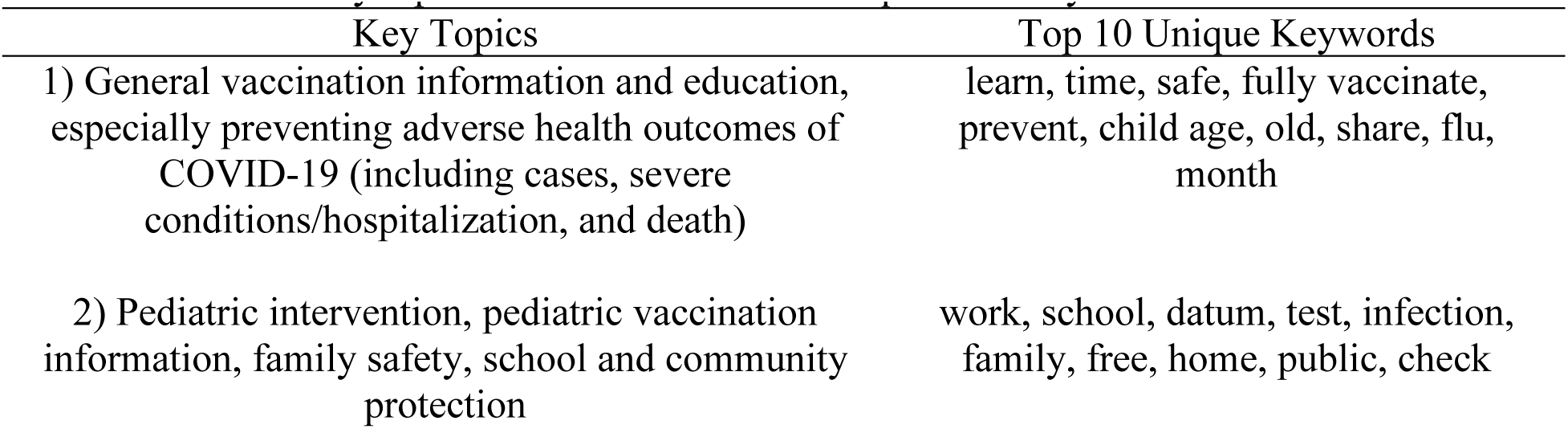
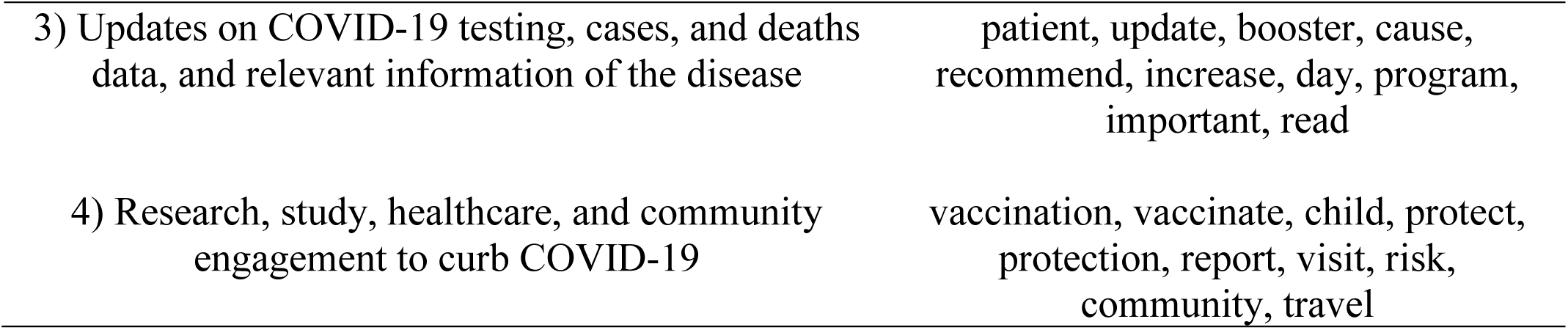
Identified key topics of CDC tweets with unique focal keywords

Visualization of the topics were displayed through an 2D plane, or an interactive chart. Based on LDA visualization results, these four identified key topics had the largest distances and minimal dimensional overlapping in the reduced two-dimensional plane. Nevertheless, from a public health perspective, CDC’s online health communication of COVID-19, the largest health emergency in the 21^st^ century, is relatively cohesive and comprehensive. CDC disseminated up-to-date, evidence-based, and accurate health information regarding COVID-19 to the public and to curb the pandemic. Therefore, the four key topics identified via LDA were not completely mutually exclusive. In addition, the 4-topic model had the balance of separation of topics from a computational perspective, and clear interpretability from a health perspective. Increasing the number of topics yields a substantial amount of topic overlapping, which was also challenging to provide explicit interpretations.

Visualization for key topics generated by LDA were shown in an interactive chart using pyLDAvis. In the interactive chart, topics were plotted as circles and displayed on the left panel; the most relevant terms, or associated keywords with their corresponding topics were displayed in frequency bars on the right panel, which showed each term’s frequency from each topic across the corpus (i.e., all CDC COVID-19 tweets sampled) [32] (see supplementary materials for more detailed information about visualizations in pyLDAvis). Size of the circle indicated prevalence of that topic in the corpus. Visualizations for all topics, displayed in circles on the left panel, and their top 15 corresponding relevant terms or associated keywords, displayed in frequency bars on the right panel, were provided in Appendix (Figure A1-A5 in Appendix).

Figure 1 below illustrated an example of topic 4. A list of associated terms of topic 4 and their overall frequency of the term in the corpus were displayed on the right panel. “booster”, “school”, “increase”, “parent”, and “country” were the five key terms from topic 4 based on overall frequency across all tweets.

### Multivariate Time Series Analysis Results

#### Cross-correlation Functions (CCF)

Time series of CDC topics and COVID-19 metrics were plotted to visually examine patterns and potential associations. A total of 16 time series plots (four topics x four COVID-19 epidemic metrics) were generated (Figure A14-A29 in Appendix). Cross correlation functions (CCF) were computed to quantify the dynamic association between each CDC key topic series and each of the four COVID-19 epidemic metrics. Quantitative results were presented (Table A3-A6 in Appendix). Visualizations (Figure A30-A44 in Appendix) illustrated CCFs between both type of time series. CCF values and plots showed that CDC’s key COVID-19 tweet topic series were not substantially correlated with confirmed COVID-19 case counts series. As an example, there were no specific patterns between topic 2 and daily confirmed COVID-19 cases (Figure 2a).

COVID-19 confirmed cases and death time series had very similar dynamic patterns in the U.S. across time span (Figure 2b). Consequently, they also showed similar CCF with CDC key topic series (Figure A45 in Appendix). COVID-19 deaths had no substantial correlations with any of the four CDC key topics (Figure A18-A21 in Appendix) based on CCFs. There were no substantial correlations between any of the four key topics and COVID-19 testing series, as well as fully vaccinated rate series. Examples showed the CCFs between those and topic 2 (Figure 3-4). These results indicated potential discrepancy between CDC’s health communication focus and actual COVID-19 epidemic dynamics in the U.S. during the pandemic.

**Figure 3.**
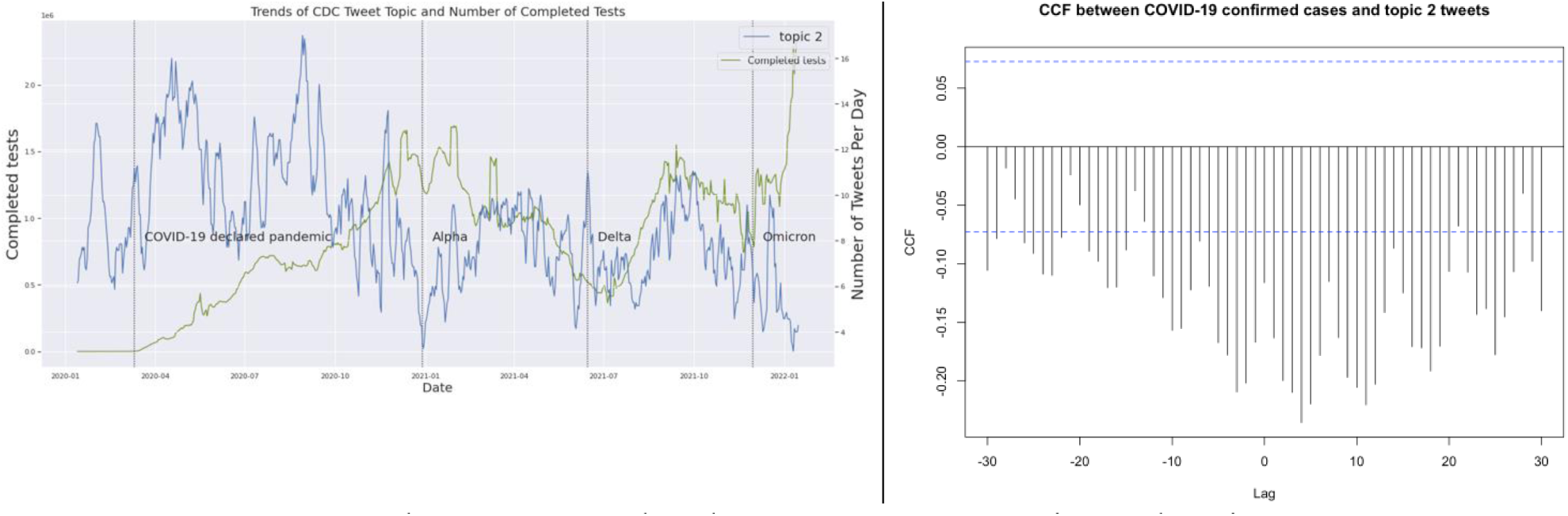
CCF between completed COVID-19 tests series and topic 2 tweets

**Figure 4.**
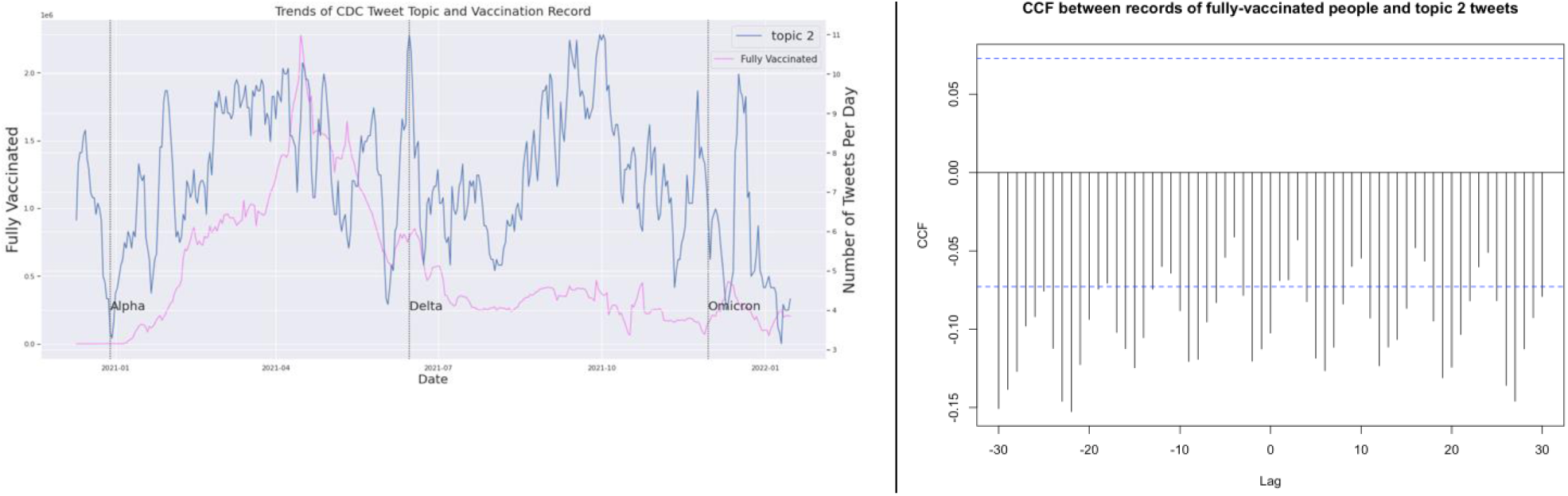
CCF between completed COVID-19 vaccinations series and topic 2 tweets

#### Mutual Information (MI)

Mutual information (MI) between each CDC tweet topic series and each COVID-19 metric series were calculated and shown in Table 4. Confirmed case counts and topic 4 (research, healthcare, and community engagement to restrain COVID-19) had the highest MI value (3.21), indicating that there is a large dependency in COVID-19 case and topic 4. On the other hand, vaccinate rate series and topic 3 had the lowest MI (0.56), indicating almost independence between the two series. Among all four key topics, topic 4 showed the highest MI (3.21, 3.02, 3.21, 1.65) with the four COVID-19 metrics series. Topic 2 (pediatric intervention, family safety, school, and community protection) had smaller MI values with the COVID-19 metric series than topic 4. MI of topic 1 (information on COVID-19 vaccination and education on preventing its adverse health outcomes) and topic 3 (updates on COVID-19 testing, cases, and deaths metrics, and relevant information of the disease) showed similar values with all four COVID-19 metrics series overall, though MI values of topic 1 were slightly higher. Vaccination and educational information on adverse health outcomes of COVID-19 appeared not to substantially be correlated with COVID-19 epidemic metrics, including testing, cases, and death. We speculated that CDC considered vaccination and preventing adverse health outcomes of COVID-19 both critical to public health and disseminated these topics regardless of current COVID-19 situation.

**Table 4.**
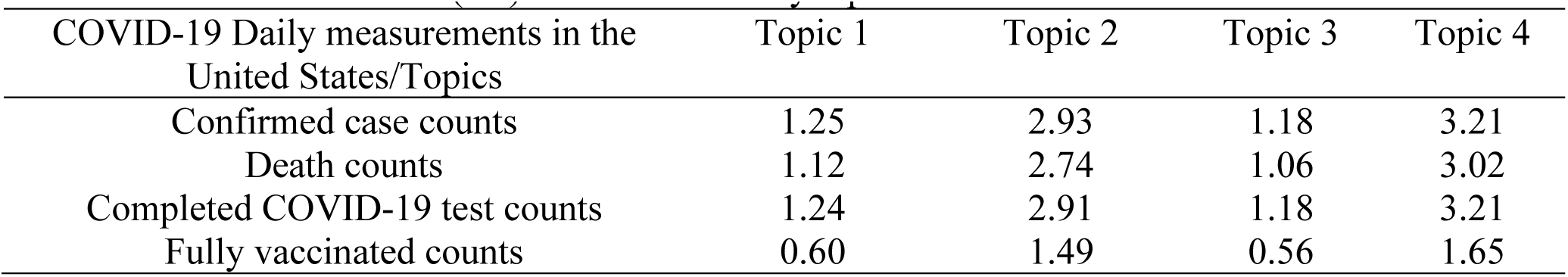
Mutual information (MI) between CDC key topic and COVID-19 metrics in the U.S.

In addition, MIs between all pairs of CDC topic time series were calculated (Table A7 in Appendix). The resulting MIs, ranked from largest to smallest, was topic 2 and 4, topic 3 and 4, topic 1 and 2, topic 2 and 3, topic 1 and 4, and topic 1 and 3. Based on CDC’s COVID-19 tweeting pattern, pediatric intervention, family and community safety were strongly associated with healthcare research studies and public engagement to curb the spread of COVID-19.

#### Autoregressive Integrated Moving Average with External Variable (ARIMAX)

We reported ARIMAX performance measures, including values of ARIMAX parameters (*p*, *d*, *q*), dAIC (difference in Akaike Information Criterion), RMSE (Root-Mean-Squared error), and MAE (Mean Absolute Error) in Table 5 below. As an external input, vaccination rate time series significantly improved the performances of the original ARIMA models for all CDC key topics (dAIC = -108.15, -69.79, -90.54, -91.53 for topic 1 to 4, respectively). This was the largest increase in model performance across all topics as exogeneous variable in ARIMAX model. COVID-19 case series improved the ARIMA model performances for CDC topic 1 and 3 (dAIC = -104.76, -1.53 for topic 1 and 3, respectively). Including death or testing series did not show improvement to the ARIMA model performances for all CDC key topics.

**Table 5.**
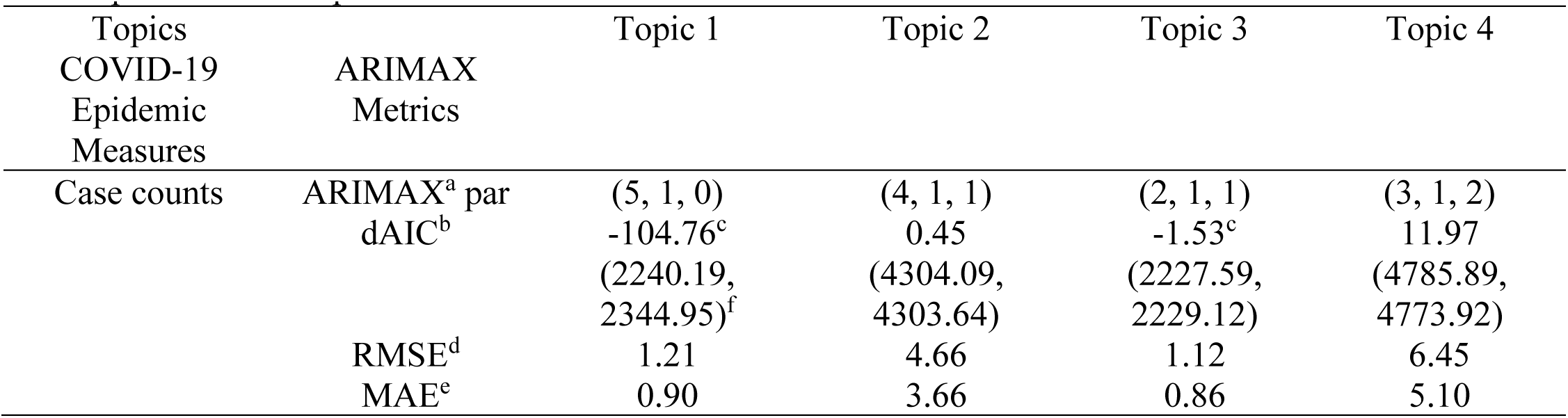
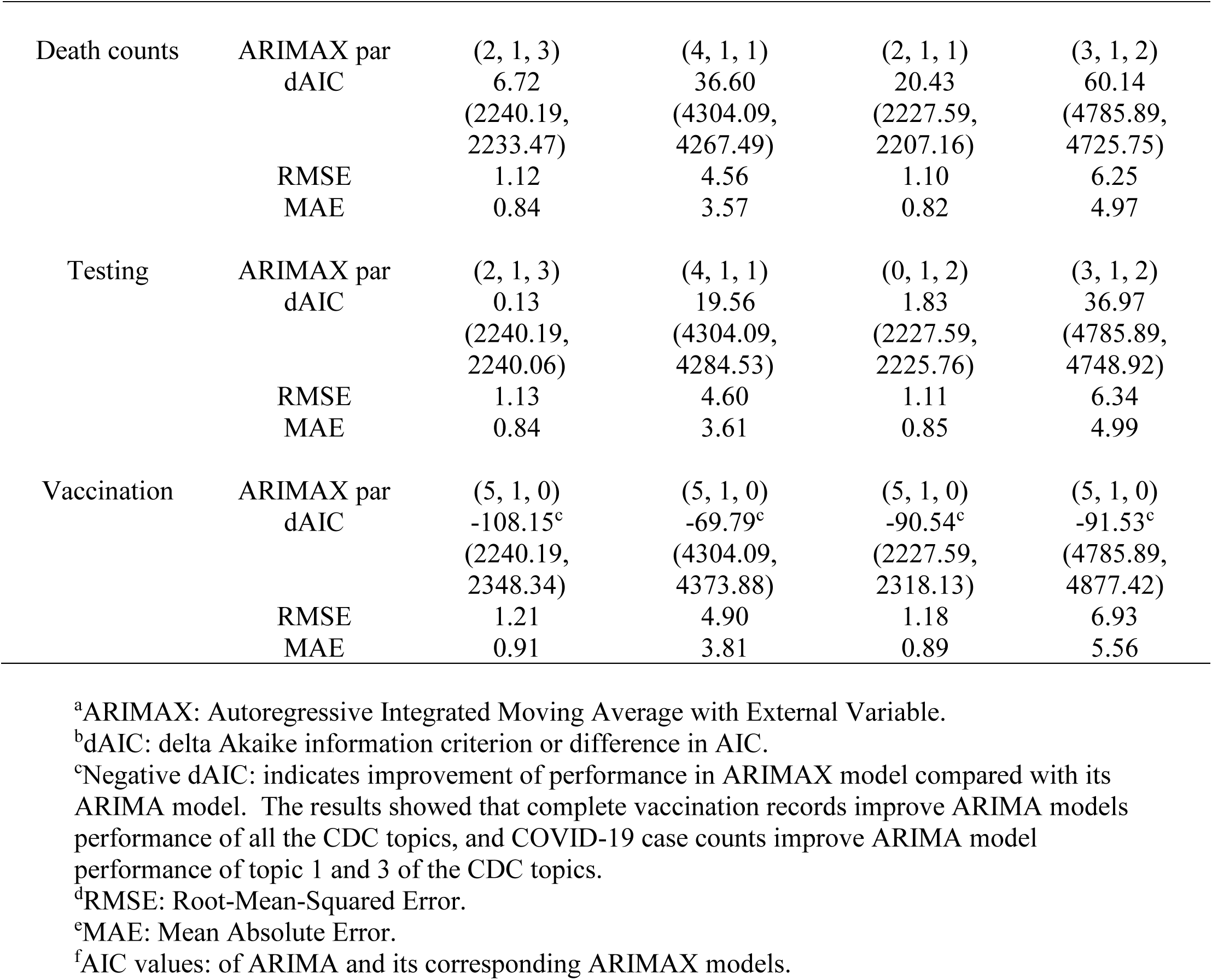
ARIMAX parameters, AIC, RMSE, and MAE of each CDC topic and COVID-19 epidemic metric pair

ARIMAX models with lower RMSE and MAE values indicate higher accuracy of the time series models (Table 5 below). Overall, ARIMAX models for topic 1 and topic 3 with all COVID-19 measures delivered the smallest RMSE values (lowest 1.10 for topic 3 with death counts, highest 1.21 for topic 1 with full vaccination records), while those of topic 4 delivered the largest RMSE values (lowest 6.25 with death counts, highest 6.93 with full vaccination records). Similarly, MAE values were the lowest for ARIMAX models for topic 1 and topic 3 with the epidemic metrics (lowest 0.82 for topic 3 with death counts, highest 0.91 for topic 1 with full vaccination records), and they were the largest for topic 4 with the epidemic metrics (lowest 4.97 with death counts, highest 5.56 with full vaccination records). These ARIMAX performance results showed significant variabilities between the two types of time series (CDC key topics and COVID-19 metrics in the U.S.).

We performed exhaustive search to identify the optimal ARIMAX parameters (*p, d, q*). For example, topic 1 with death counts and completed testing records have the same parameter values (*p*, *d*, *q* = 2, 1, 3), indicating that the optimal ARIMAX model between these time series needed first-degree differencing (*d* = 1) to achieve stationarity and minimal AIC value, its autoregressive time lag is 2 (*p* = 2), and its moving average time lag is 3 (*q* = 3). Topic 1 series with case counts and complete vaccination records have the same parameter values (*p*, *d*, *q* = 5, 1, 0), indicating that the model is simply an autoregressive model (*q*=0 with no moving average terms) with time lag of 5 (*p* = 5) after differencing once (*d* = 1). The complete ARIMAX parameters were shown in Table 5. All ARIMAX models needed first degree differencing (*d* = 1) to be stationary and to minimize AIC values.

## Discussion

### Principal Findings

Our study took a unique approach of infoveillance by identifying potential associations between COVID-19 epidemic outcome metrics in the U.S. and CDC’s key topic dynamics during different stages of the pandemic. This innovative framework significantly expanded the original infoveillance approach that generally relied on number of posts (i.e., posting dynamics) without further extracting more detailed and meaningful content topics and sentiments from the textual data. Our study was able to further provide practical and useful health communication strategies for public health agencies to effectively communicate to the public with timely and accurate information. We systematically investigated and comprehensively identified CDC’s key topics, COVID-19 epidemic outcome metrics, and dynamic associations between the two types of data series based on more than 1.7 million COVID-related English tweets from CDC since January 2022. LDA topic model was built to characterize and identify the dynamic shifts of topics in CDC’s COVID-19 communication over more than 2-year period. For the first time, we were able to identify the four key topics: 1) general vaccination information and education; 2) pediatric intervention that also involved family and school safety; 3) updates on the COVID-19 epidemic situation such as numbers of cases, deaths, etc.; 4) research studies that was able to curb the pandemic.

Our study is the first of the kind to comprehensively evaluate the impact of online communications, especially on Twitter, one of the major social media platforms, during different phases of a large health emergency. We used NLP, especially topic modeling to explicitly explore CDC’s COVID-19 communication on social media. This approach and findings significantly reduced the gap of previous studies that overlooked the dynamic association between detailed topics discussed by public health agencies on social media and the real-world epidemic outcome metrics. We examined the dynamic associations between the four identified key topics, and four COVID-19 epidemic outcome metrics. Among the four major topics, topic 1, which covered information on vaccination and adverse health outcomes of COVID-19, had substantially strong associations with death counts and testing records during the Alpha phase (December 29, 2020 – June 14, 2021). Topic 3, which provided updates on three of the COVID- 19 measures (testing, cases, and deaths) and their relevant information, aligned better with case series during the Delta phase (June 15, 2021 – November 29, 2021). It also matched with death series during the declared pandemic phase (original variant, March 11, 2020 – December 28, 2020) and Delta phase, classified by WHO on May 11, 2021. Furthermore, even though topic 3 did not demonstrate visible temporal association with the testing series, timely communication from CDC was actually strongly associated with the trend over the entire study period based on the multivariate time series analysis.

According to these findings of this study, we suggest that aligning the content topics of health communication from public health agencies with the temporal dynamics of COVID-19, or other public health emergencies (e.g., major epidemic outcome metrics) can help health agencies provide more timely and relevant information to the public. Therefore, we recommend that CDC and other public health agencies to monitor the epidemic outcome metrics in real-time. Health agencies can post timely updates about the emergency, most recent findings, and interventions on social media according to the dynamic changes of these outcome metrics. Public health agencies can regain trust from the public by not only helping the public better understand the complex dynamics of the health emergency, but also informing the public with evidence-based guidance and recommendations more effectively.

## Limitations and Future Work

There are several limitations in this infoveillance study that could be improved in future work. First, while we focused on probabilistic-based LDA for topic modeling, there are other alternative natural language processing (NLP) options such as deep learning-based bidirectional encoder representations from transformers (BERT). Hence, we will explore BERT and other state-of-the-art NLP techniques for content topic modeling and sentiment analysis in the future. Second, public engagement (i.e., retweets, likes, and replies, etc.) of CDC’s health communication is an important indicator of the effectiveness of health agencies’ communication efforts. There have been studies that analyzed public sentiments and attitudes ([9]–[12]) towards various health-related topics. However, very few investigated the associations of public sentiments shifts along disease-related metrics. In addition, public sentiments and attitudes are heavily influenced by health agencies’ messages and should not be misled by misinformation and fake news. Public opinions influence health practices and interventions, which make significant impact on the actual epidemic trends. Thus, it is important to discover the underlying association between public health communication topics and actual epidemic measures. The insights can help public health agencies develop better social media strategies to address public concerns. Therefore, we suggest that further examining the dynamics and patterns of public responses to CDC’s original tweets can gain valuable insight on public perceptions and attitudes around various issues during the pandemic, such as pharmaceutical interventions (e.g., vaccination) and non-pharmaceutical interventions. For example, we could conduct content analysis to explicitly identify public concerns towards CDC’s health communications. In addition, sentiment analysis could also be applied to extract public sentiments (i.e., positive, neutral, or negative) towards CDC’s health communications, and further help identify public attitudes and reactions to various content topics that CDC has communicated. Public attitudes will ultimately determine individual health behavior and decision-making, such as vaccination acceptance and compliance with NPIs, which in turn drive the overall epidemic dynamics.

Therefore, it is critical to investigate real-time public engagement, such as retweeting or replying on social media towards public health agencies’ health communications, to better inform health agencies to prioritize their communications and address public concerns about specific content topics.

### Conclusions

This study investigated the dynamic associations between CDC’s detailed COVID-19 communication topics on Twitter and epidemic metrics for almost two years during the pandemic. Using LDA topic modeling, we were the first to comprehensively identify and explore various COVID-related topics tweeted by the federal public health agency during the pandemic. We also collected daily COVID-19 epidemic metrics (confirmed case counts, death counts, completed tests records, and fully vaccinated records) and performed various multivariate time series analysis to unravel the temporal patterns and associations with CDC’s COVID-19 communication patterns, i.e., investigating the dynamic associations between the time series of each topic generated by LDA model and the time series of each epidemic metrics. The results suggested that some topics were strongly associated with certain COVID-19 epidemic metrics, indicating that advanced social media analytics (e.g., natural language processing, NLP) could be a valuable tool for effective infoveillance. Based on our findings, we recommend that CDC, along with other public health agencies, could further optimize their health communications on social media platforms by posting contents and topics that aligns with the temporal dynamics of key epidemic metrics. Effectively disseminating timely and accurate information about an ongoing health emergency to the general public is crucial for health agencies to inform and educate the public, and eventually curb the ongoing health emergency. For example, we suggest increasing online health communications on health practices and interventions during high-level epidemic periods with large numbers of cases and deaths. Our findings also highlighted the importance of health communications on social media platforms to better respond to and tackle future health emergencies and issues.

## Supporting information

Multimedia Appendix

## Data Availability

All data produced in the present study are available upon reasonable request to the authors

## Acknowledgements

We thank Naomi Nikita Thammadi, former graduate student of University of North Carolina at Charlotte, who helped with data collection through Twitter API and initial data preprocessing. This project was partially supported by the Models of Infectious Disease Agent Study (MIDAS) Network grant MIDASUP-05 to SC.

## Abbreviations

AIC: Akaike Information Criteria
ARIMA: string-nameoregressive Integrated Moving Average
ARIMAX: string-nameoregressive Integrated Moving Average with External Variable CCF: cross-correlation functions
CDC: Centers for Disease Control and Prevention dAIC: difference in Akaike information criterion LDA: latent Dirichlet allocation
MAE: Mean Absolute Error MI: mutual information
RMSE: Root-Mean-Squared Error

