## Supplementary material for "Infoveillance study on the dynamic associations between CDC social media contents and epidemic measures during COVID-19": Multimedia Appendix

### Supplementary materials

Latent Dirichlet allocation (LDA) topic model:

- A corpus is denoted by D = {$\mathbf{w_1}$, $\mathbf{w_2}$, …, $\mathbf{w_M}$}, where M = the number of documents within the corpus
- Each topic for a document is denoted by T = {$t_1$, $t_2$, …, $t_K$}, where K = the number of topics in the corpus
- Each document is denoted by $\mathbf{w}$ = ($w_1$, $w_2$, …, $w_N$), where N = the number of words in the document
- Each word in the document is indexed by {1,…,V}

LDA algorithm achieves the following tasks:

1. Forming an M x K matrix, where the weight $w_{m,k}$ = the association between a document $d_m$ and a topic $t_k$.
2. Forming an N x K matrix, where the weight $w_{n,k}$ = the association between a word $w_n$ and topic $t_k$.

Notations:
Dirichlet(i): Dirichlet distribution with parameter i;
Mult(i): multinomial distribution with parameter i;
$\beta_t$: word distribution for topic *t*;
$\theta_d$: topic proportion for document *d*;
$\eta$ and $\alpha$: hyperparameters of each of their Dirichlet distributions.
The respective generative processes for each task are shown below [1]:

1. For each topic t ∈ {1, …, K}, draw a distribution over words $\beta_t$ ~ Dirichlet ($\eta$): from each document randomly draw words and assign them to K topics.
2. For each document d,
    a. Draw a vector of topic proportions $\theta_d$ ~ Dirichlet ($\alpha$):
    b. For each word $w_n$ in document d, where n ∈ {1, …, N}
        i. Draw a topic assignment $\mathbf{z}_n$ ~ Mult($\theta_d$): calculate the probability of assignments to topic *t* over all documents that come from word *w*, obtains the number of documents there are in topics *t* from the word w.
        ii. Draw a word $w_n$ ~ Mult($\beta_{zn}$): calculate the probability of words in documents *d* that are assigned to topic t, obtains the number of words belong to the topic *t* given document *d*
        iii. The probability of the word *w* that belongs to topic *t* is the product of the above two probabilities

Autoregressive (AR) model:
AR is a regression model where linear combinations of the past terms are the predictors of the variable in the future. Assuming the values of the variable are independently and normally distributed, AR(*p*) model of order or lag *p* can be written as:

$$y_t = c + \phi_1 y_{t-1} + \phi_2 y_{t-2} + \ldots + \phi_p y_{t-p} + \varepsilon_t = c + \sum_{i=1}^{p} \phi_i y_{t-i} + \varepsilon_t \text{ [24]},$$

or in a simplified version with backward shift operator form:

$$\phi_p(B)y_t = 1 - \phi_1 B + \ldots + \phi_p B_p \text{ [25]},$$

where $c$ = average of changes between the sequential observations,
$\varepsilon_t$ = white noise,
$y_t$ = lagged values, or predictor values of the next value of the variable y.
MA is another linear model of past forecast errors in a univariate time series. MA($q$) model of order or lag $q$ can be written as:

$$y_t = c + \theta_1\varepsilon_{t-1} + \theta_2\varepsilon_{t-2} + \dots + \theta_q\varepsilon_{t-q} + \varepsilon_t = c + \sum_{j=1}^{q} \theta_j y_{t-j} + \varepsilon_t \text{ [26]},$$

or in backward shift operator form:

$$\theta_q(B)y_t = 1 - \theta_1 B + \dots + \theta_q B_q \text{ [25]}.$$

Autoregressive integrated moving average (ARIMA) model:
ARIMA model consists of both AR($p$) and MA($q$) as well as order $d$ differencing term, resulting in the following ARIMA ($p$, $d$, $q$) model:

$$y'_t = c + \phi_1 y'_{t-1} + \dots + \phi_p y'_{t-p} + \theta_1\varepsilon_{t-1} + \theta_2\varepsilon_{t-2} + \dots + \theta_q\varepsilon_{t-q} + \varepsilon_t \text{ [27]},$$

or in backward shift form:

$$(1 - \phi_1 B + \dots + \phi_p B_p)\nabla^d y_t = c + (1 + \theta_1 B + \dots + \theta_q B^q)\varepsilon_t \text{ or } \phi(B)\,)\nabla^d y_t = c + \theta(B)\varepsilon_t \text{ [28]}$$ (see supplementary materials for details on the parameters),

where $y'_t$ = time series after differencing, which is presented as $(1 – B)^d y_t$ as d differences in backward shift operator form,
$\nabla^d = (1 – B)^d y_t$,
$p$ = order of past lagged values of for each time point of autoregressive model,
$d$ = the degree of differencing occurred, or the number of times performs integration,
$q$ = order past lagged errors for the error term as a combination of predictors of moving average model.

Akaike information criterion (AIC):
Given a set of data, Akaike information criterion (AIC), which was proposed by Akaike in his published paper in 1974, is used as an estimator for statistical model comparison and selection. It is defined as:

$$AIC = -2\log(L) + 2k \text{ [30]},$$

where $k$ = the number of independent parameters, L = the maximum likelihood for a statistical model. AIC is widely used to estimate the model's fit, especially for time series data, greater penalties are provided accordingly to improve the fit, but could meet its limitation when data size becomes large, leading overfitting of the model. We have performed series of exhaustive searches were executed amongst all possible ARIMA models in this study for model comparison and selection.

Definition of a complete series of vaccine: either one dose of a single-dose vaccine (Johnson & Johnson viral vector vaccine) or two-dose vaccines (Pfizer or Moderna mRNA vaccine).

### Appendix

**Time span for data collection:**
Start date: December 7, 2019. End date: January 15, 2022

**Search query used to obtain CDC tweets for data collection**:
(ncov OR ncov-19 OR sars OR SARS-COV-2 OR "corona virus" OR pandemic OR pheic OR "wuhan virus" OR "china virus" OR "wuhan pneumonia" OR "wuhan flue" OR kungflue OR covid19 OR covid OR "covid 19" OR coronavirus OR vaccine OR vaccines OR vaccination)

**Stopwords list used in LDA topic model**:
{'covid','continue','good', 'include','mmwr','feel','etc','ever','sure','covid19', 'can','especially', 'many', 'well', 'our','sarscov2', 'ncov19', 'ncovcoronavirus', 'sars', 'pandemic', 'get', 'cdc', 'use', 'see', 'take', 'give', 'thank', 'still', 'show', 'first', 'keep', 'go', 'know', 'also', 'make', 'today', 'find', 'need', 'show', 'vaccine', 'health', 'amp', 'mask', 'outbreak', 'case', 'ppe', 'n95', 'shortage', 'frontline', 'new', 'health', 'help', 'people', 'year'}

The pyLDAvis visualizations are displayed in two left and right panels. Visualization features are displayed on left panel, where topics are presented, and right panels, where the associated keywords with their corresponding topics are displayed in frequency bars:
1. Circles of topics generated by the LDA topic model: displayed on the left panel in different sizes, “intertopic distance map”, showing how far or near they are from each other color-coded in blue when not selected. The color of the circle is blue when no topic is being selected and is red when selected. The size indicates the prevalence of that topic in the corpus.
2. List of associated keywords of the topics in the corpus
3. Saliency (blue bars): the overall frequency of a word in the corpus, shown when no word is being selected.
4. Relevance (red bars): shows how much information a word explains about a topic by measuring the frequency of a word within a topic with weight λ.

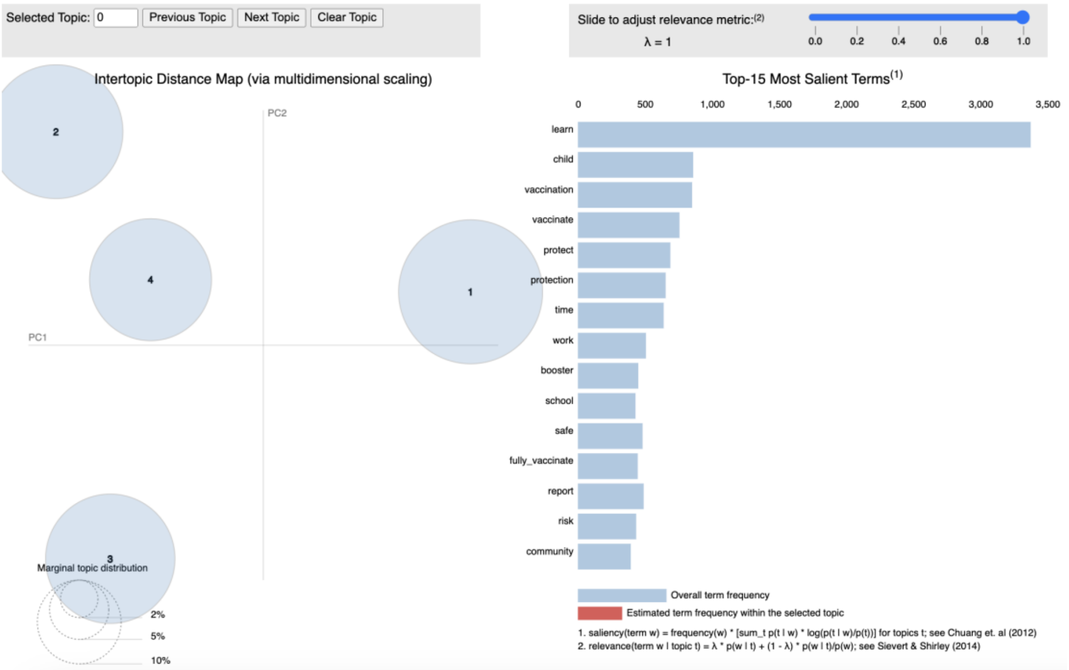

Figure 1. Interactive mapping of the topics generated by LDA

Topic 1 and its associated keywords/relevant terms:

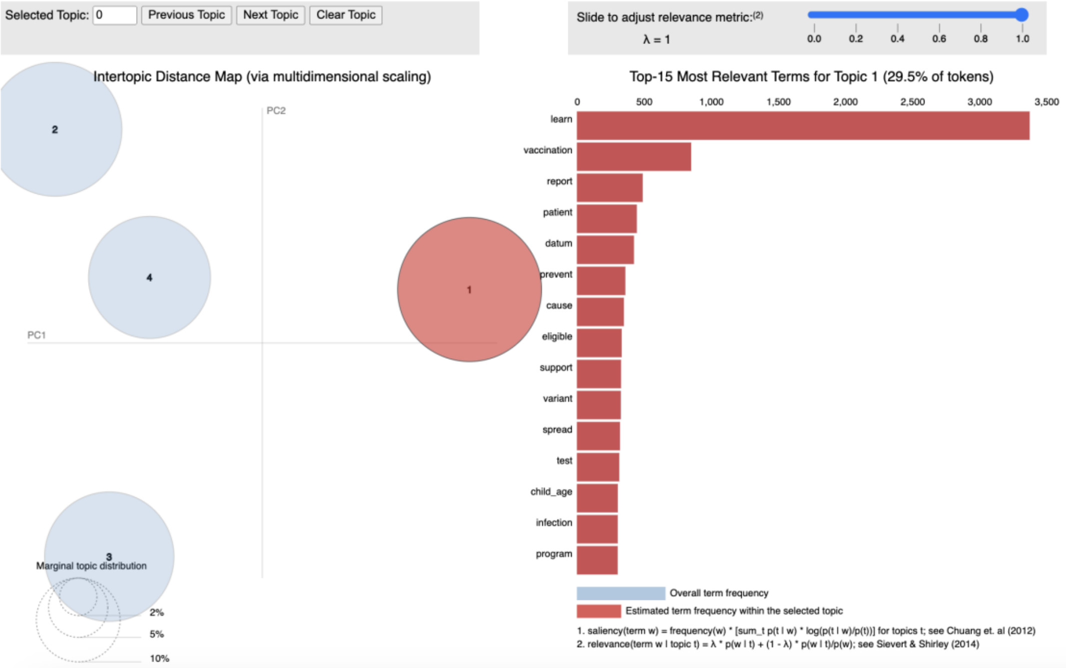


Figure 2. Interactive mapping of topic 1 generated by LDA

Topic 2 and its associated keywords/relevant terms:

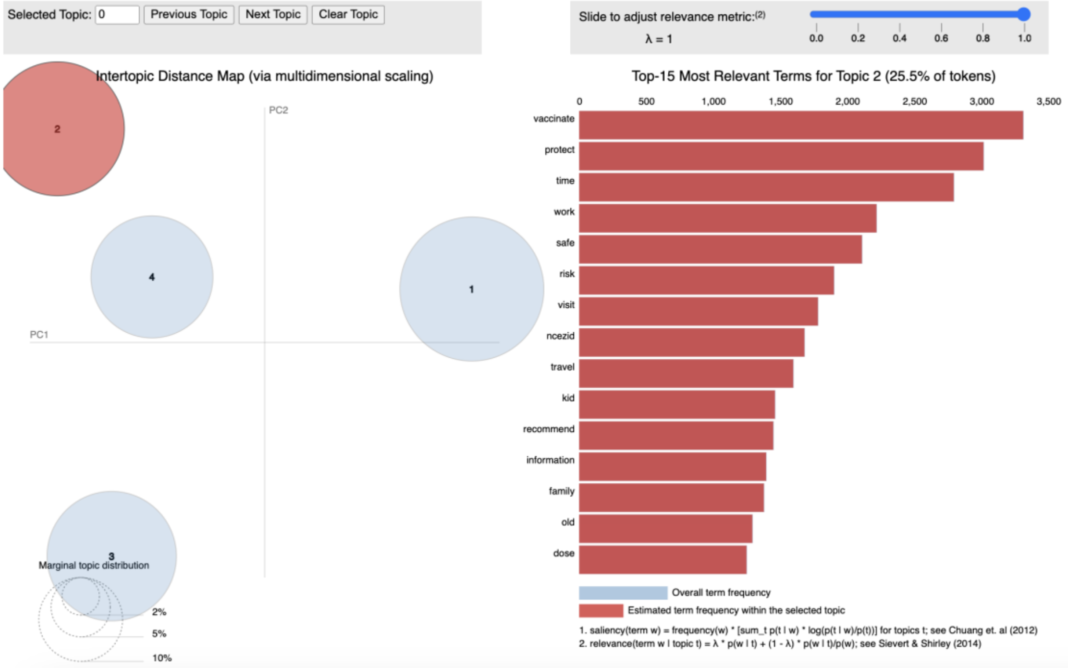


Figure 3. Interactive mapping of topic 2 generated by LDA

Topic 3 and its associated keywords/relevant terms:

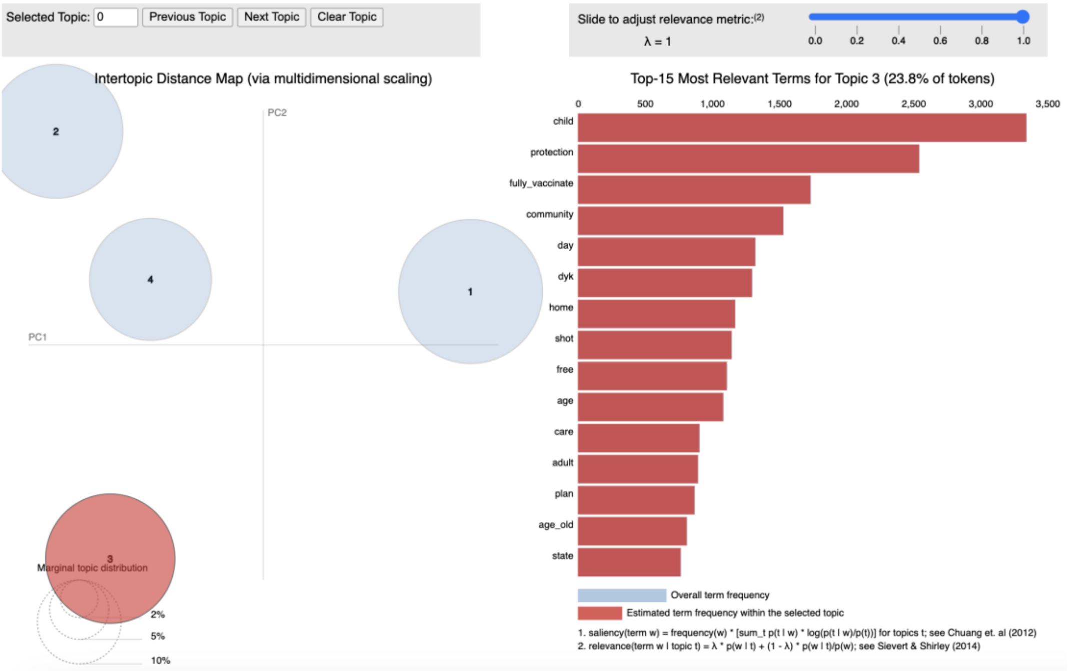


Figure 4. Interactive mapping of topic 3 generated by LDA

Topic 4 and its associated keywords/relevant terms:

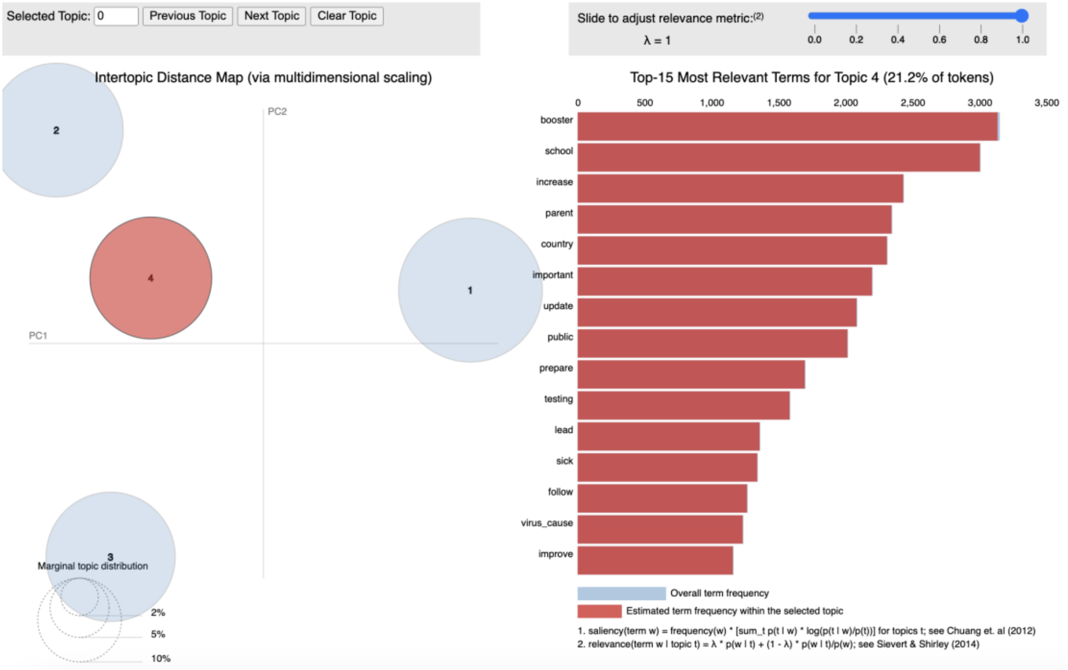


Figure 5. Interactive mapping of topic 4 generated by LDA

**Table 1**. Identified prominent topics of CDC tweets and their respective unique focal keywords

| Topic Theme | Top 10 Unique Keywords |
|---|---|
| 1) General vaccination information and education, especially preventing adverse health outcomes of COVID-19 (including cases, severe conditions/hospitalization, and death) | learn, time, safe, fully vaccinate, prevent, child age, old, share, flu, month |
| 2) Pediatric intervention, pediatric vaccination information, family safety, school and community protection | work, school, datum, test, infection, family, free, home, public, check |
| 3) Updates on COVID-19 testing, cases, and deaths data, and relevant information of the disease | patient, update, booster, cause, recommend, increase, day, program, important, read |
| 4) Research, study, healthcare, and community engagement to curb COVID-19 | vaccination, vaccinate, child, protect, protection, report, visit, risk, community, travel |

**Individual time series plots: the four COVID-19 epidemic measurements and the four CDC topics after detrending and testing for stationarity.**

X-axis shows the number of days, and y-axis shows the number of the measurement on daily basis (in blue), 7-day moving average (in red), and standard deviations (in green):

- Confirmed cases:

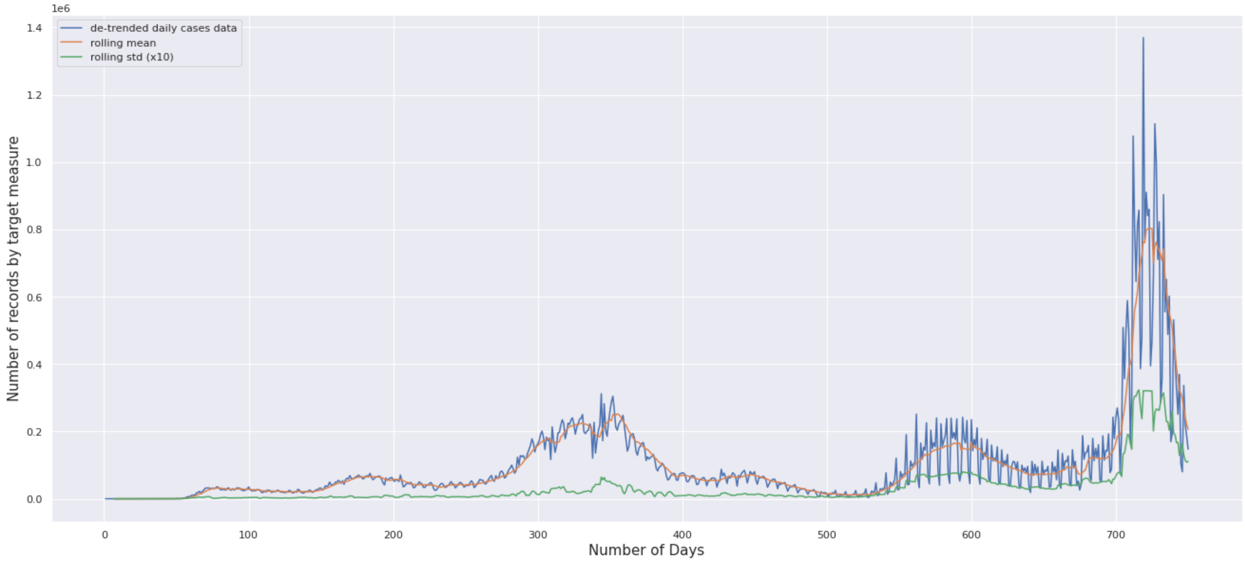


Figure 6.

- Deaths:

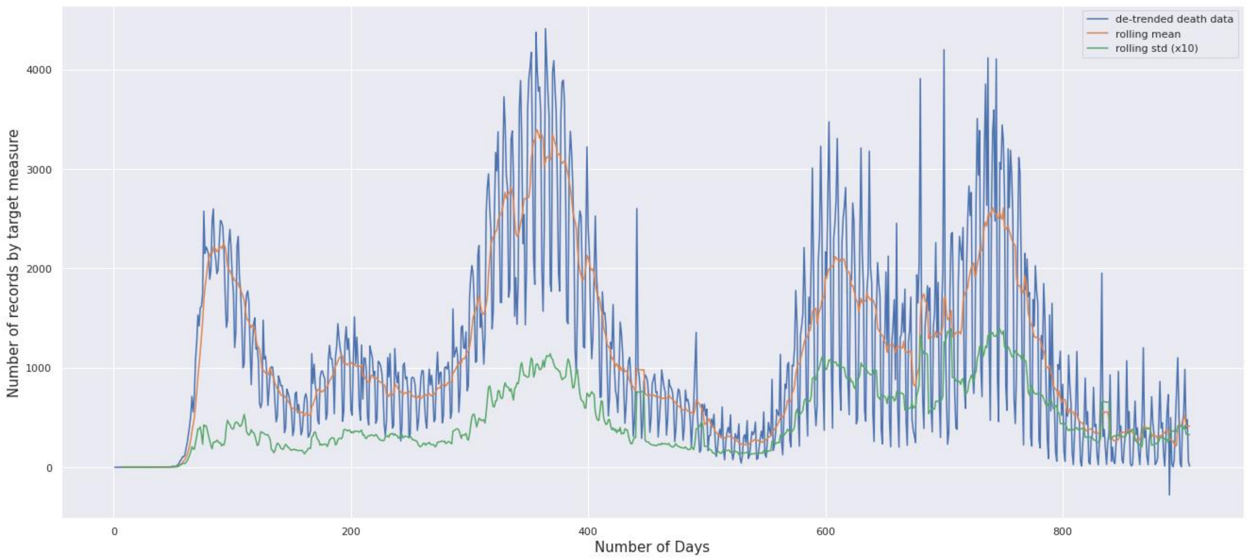


Figure 7.

- Testing:

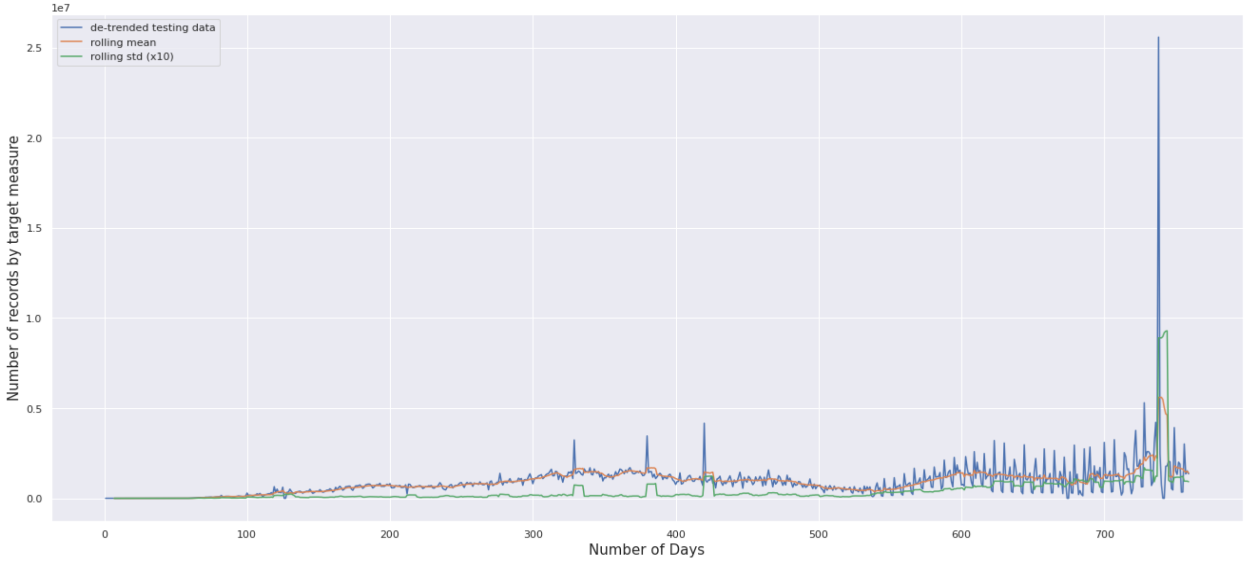


Figure 8.

- Vaccination:

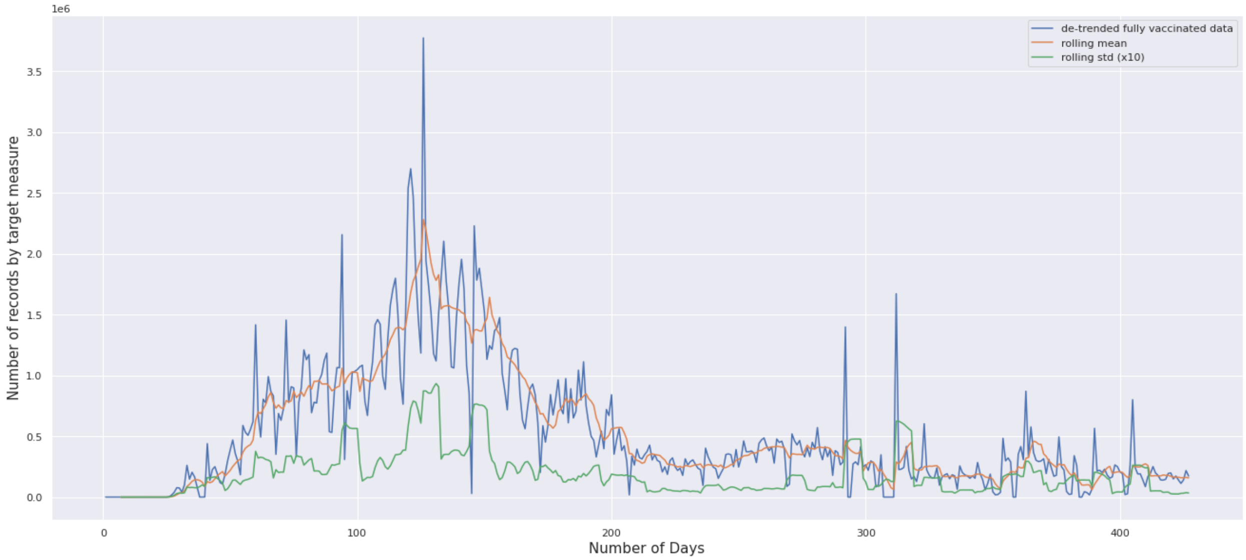


Figure 9.

- Topic 1: General vaccination information and education, especially preventing adverse health outcomes of COVID-19 (including cases, severe conditions/hospitalization, and death)

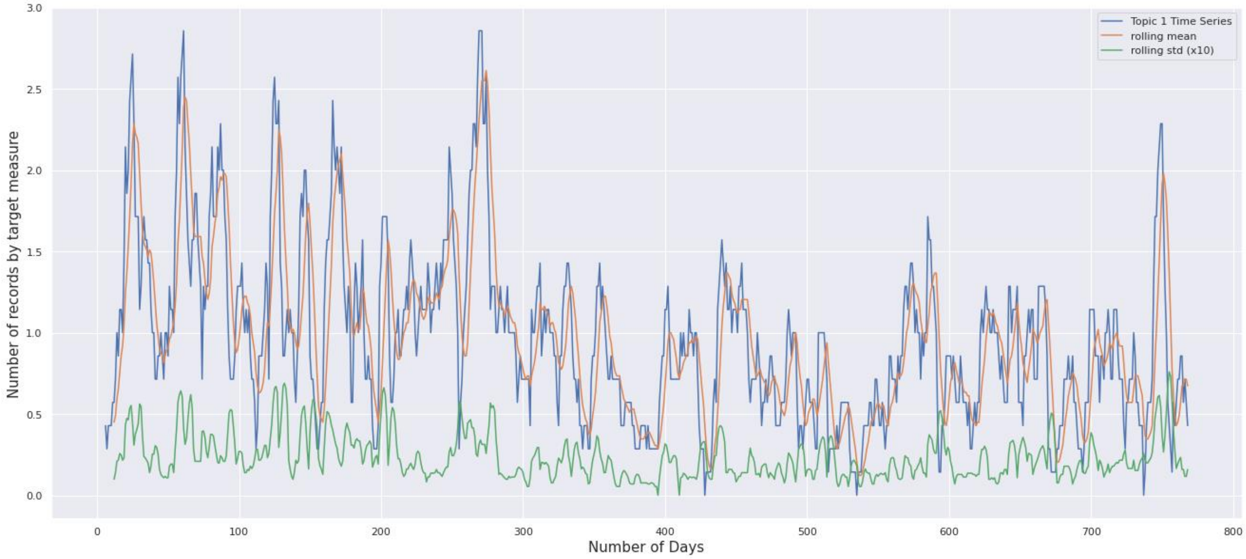


Figure 10.

- Topic 2: Pediatric intervention, pediatric vaccination information, family safety, school and community protection

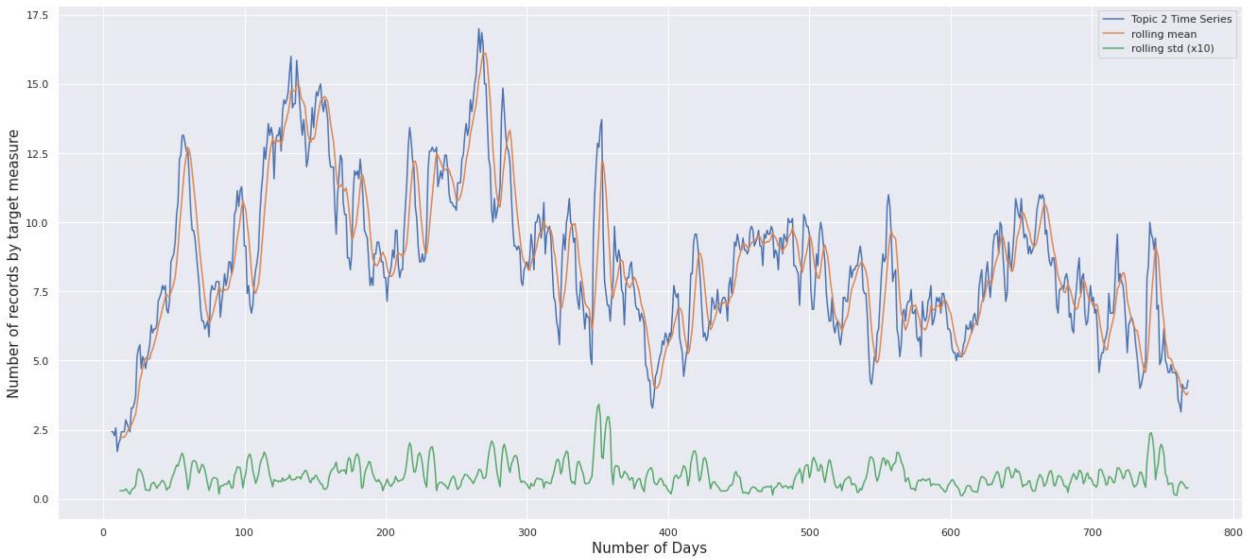


Figure 11.

- Topic 3: Updates on COVID-19 testing, cases, and deaths data, and relevant information of the disease

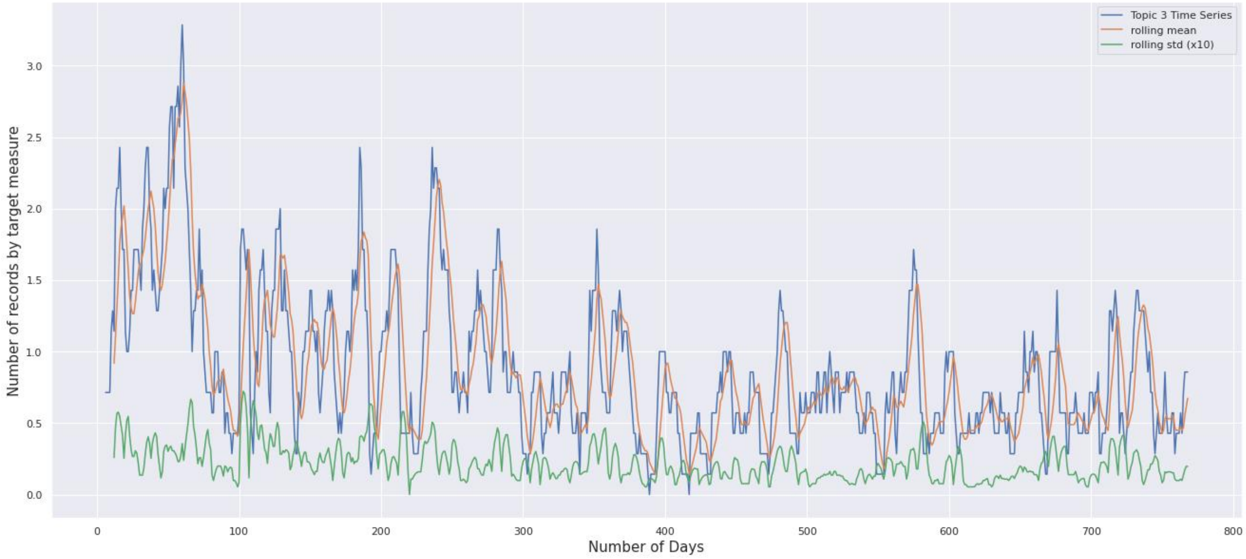


Figure 12.

- Topic 4: Research, study, healthcare, and community engagement to curb COVID-19

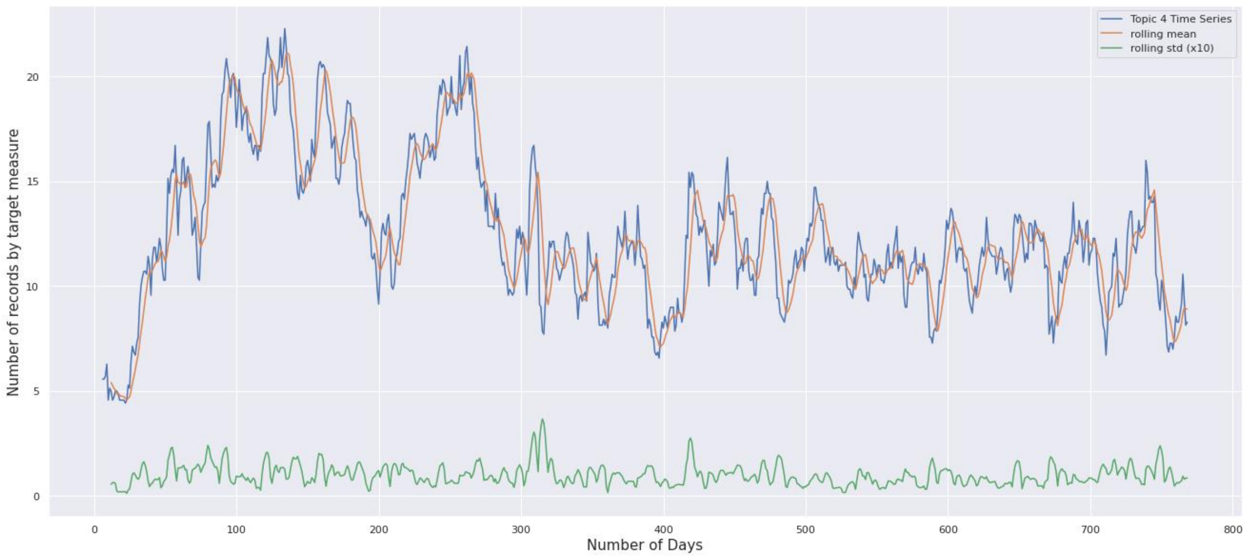


Figure 13.

#### Time series plots: CDC topics and COVID-19 epidemic measurements in dual axis

- Cases and Topic 1:

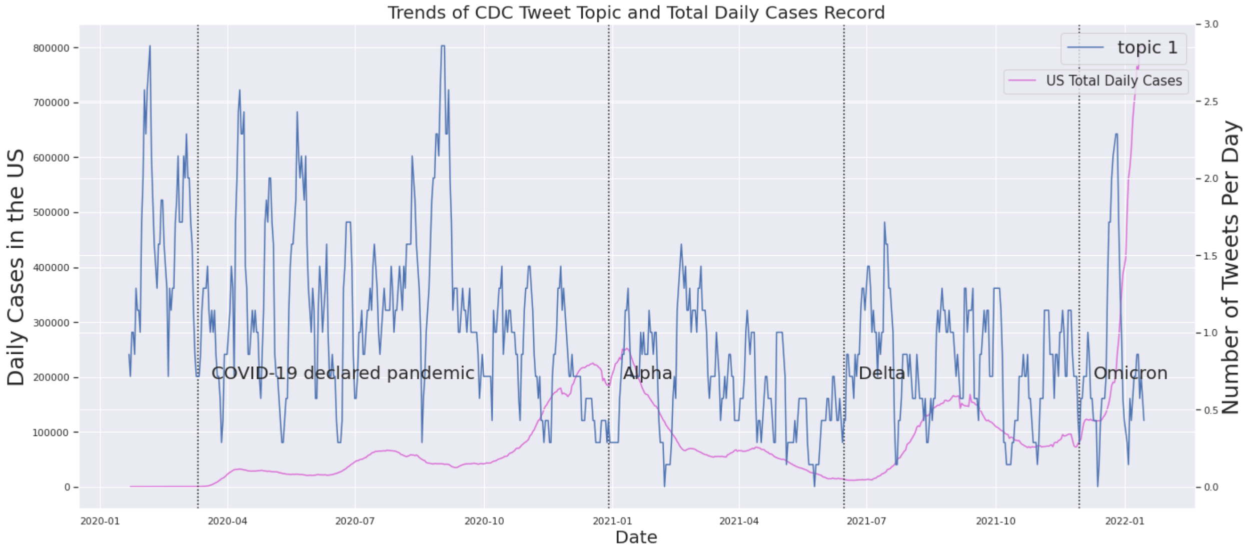


Figure 14.

- Cases and Topic 2:

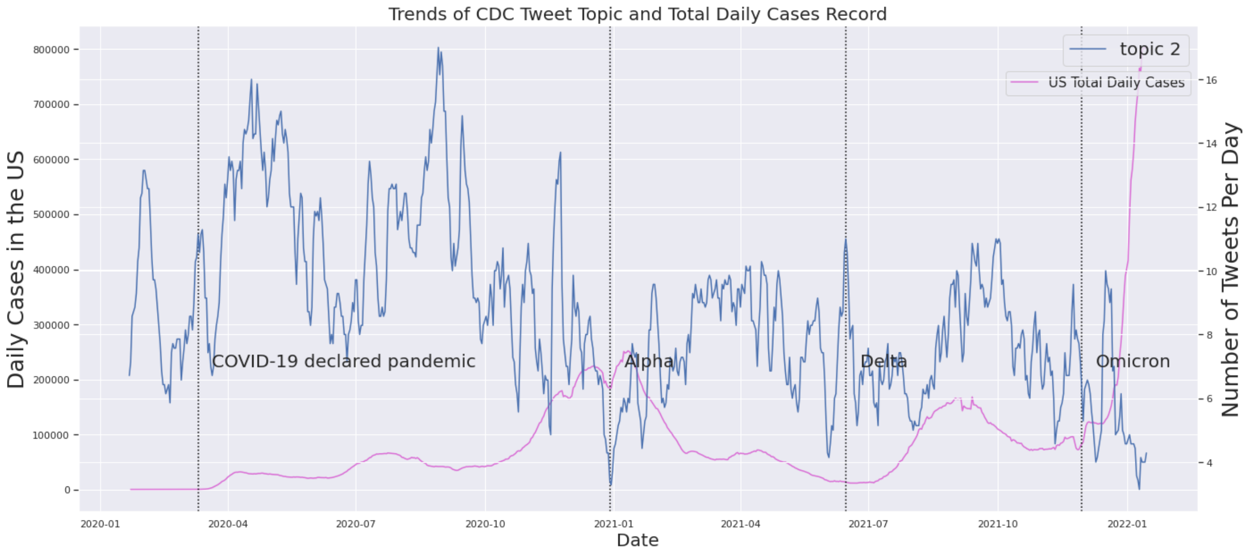


Figure 15.

- Cases and Topic 3:

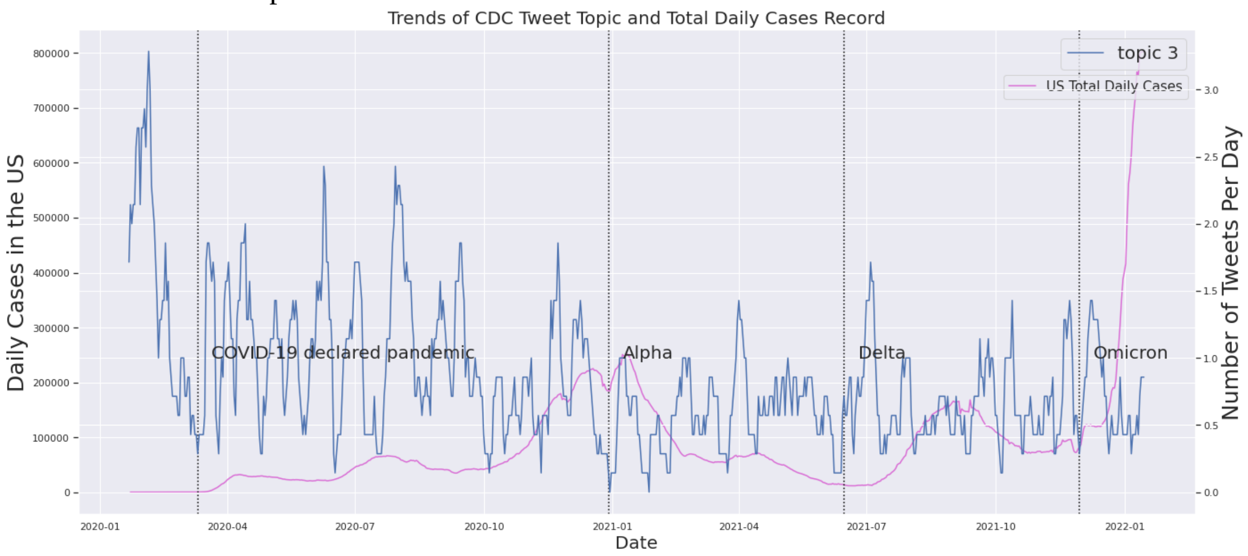


Figure 16.

- Cases and Topic 4:

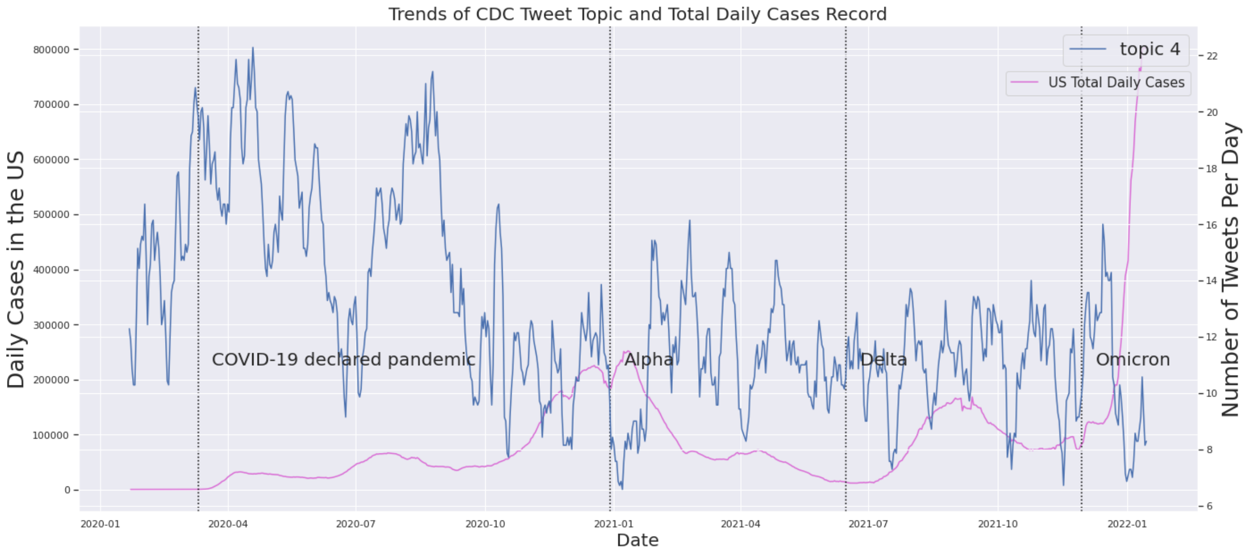


Figure 17.

- Deaths and Topic 1:

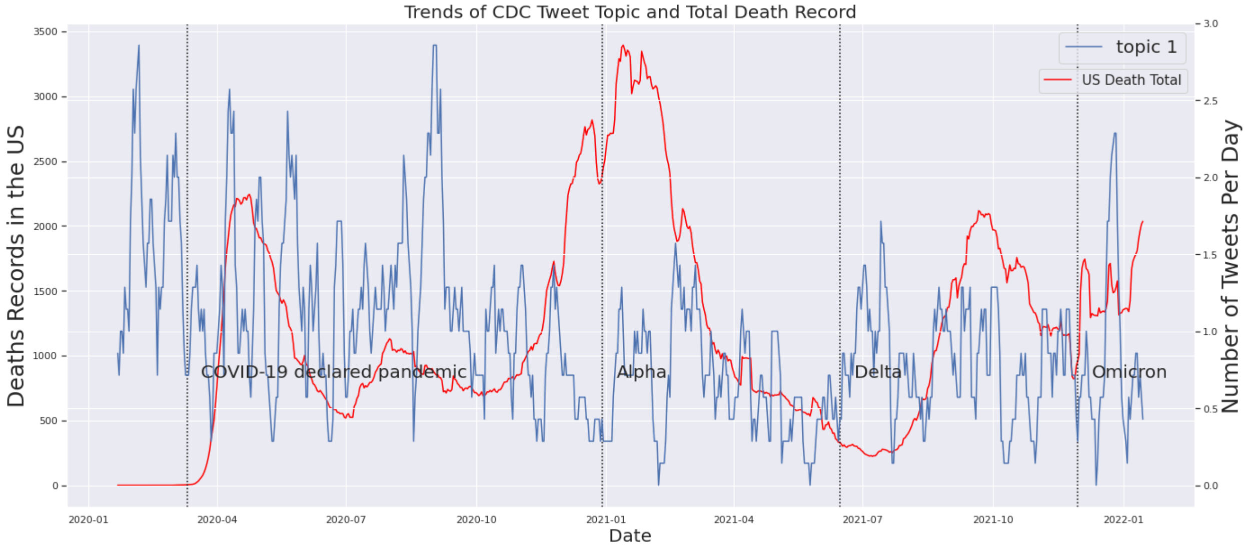


Figure 18.

- Deaths and Topic 2:

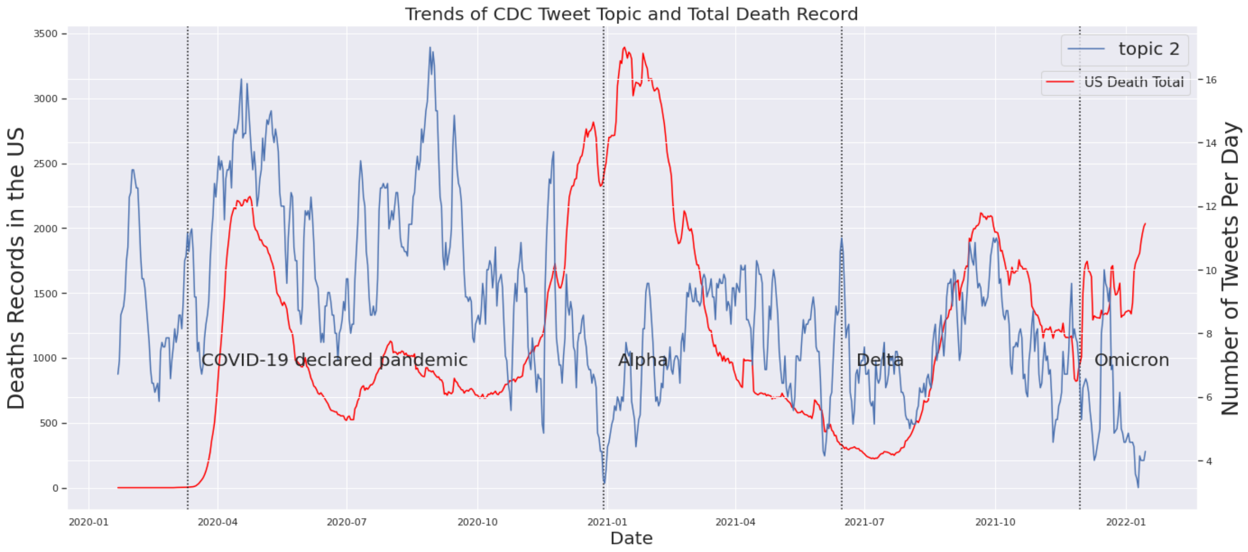


Figure 19.

- Deaths and Topic 3:

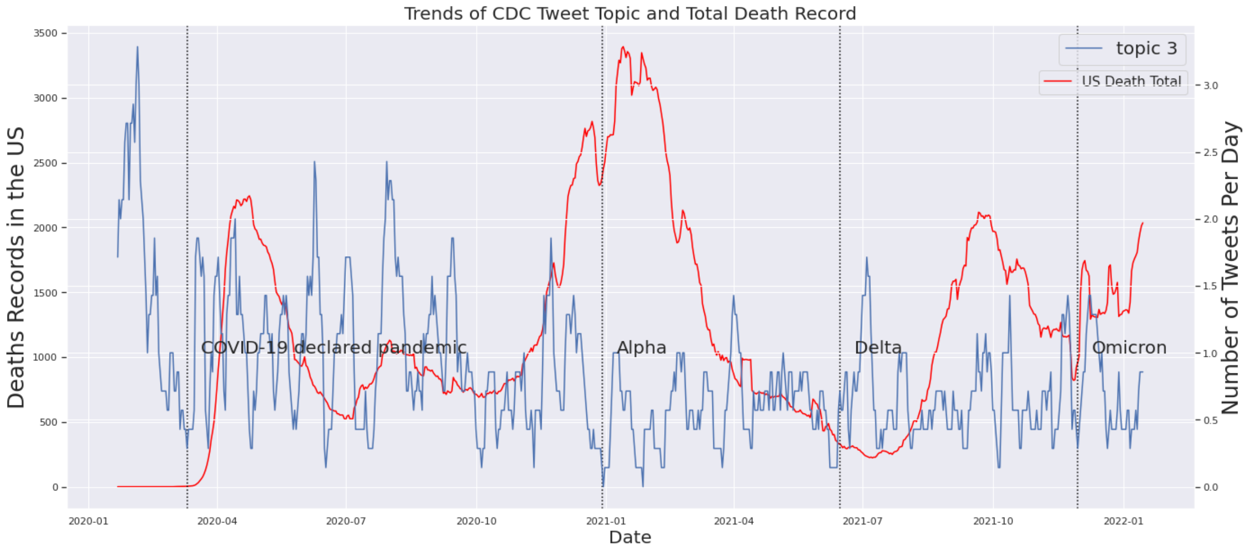


Figure 20.

- Deaths and Topic 4:

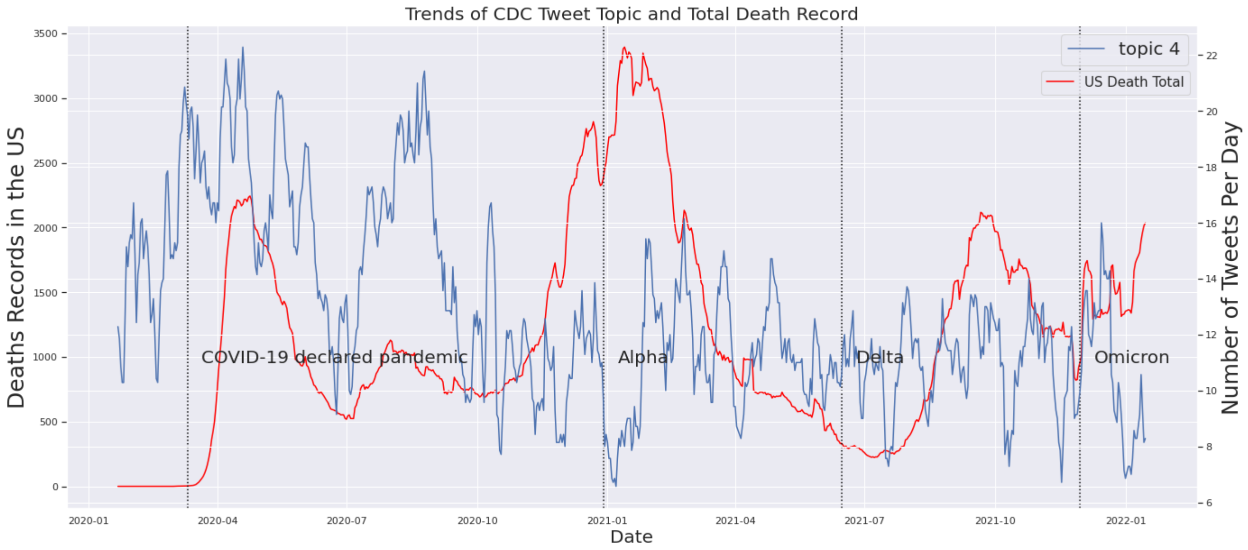


Figure 21.

- Tests and Topic 1:

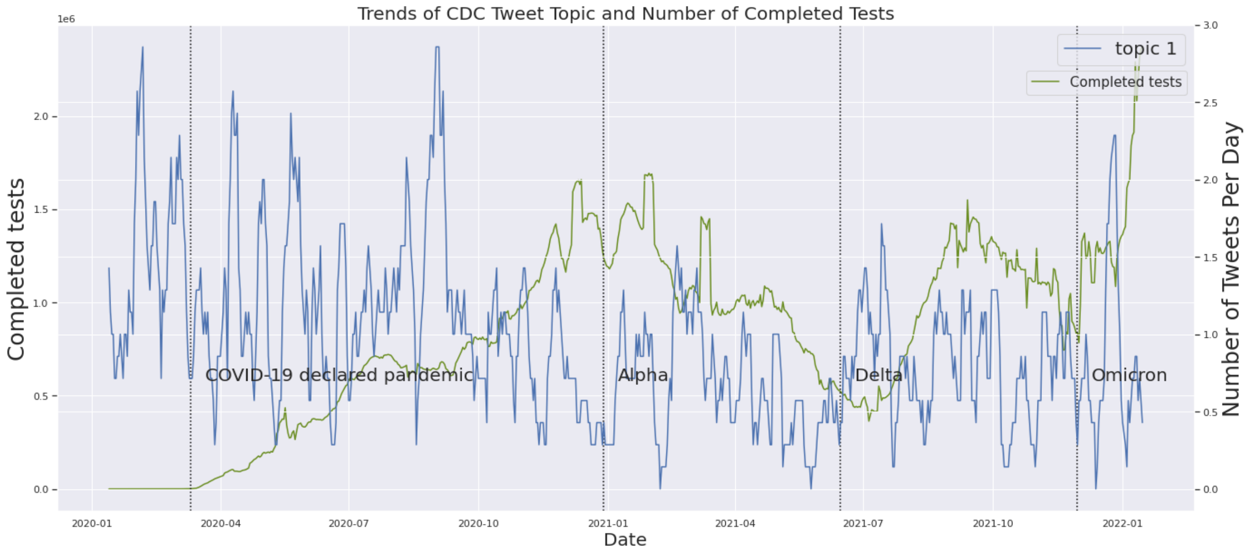


Figure 22.

- Tests and Topic 2:

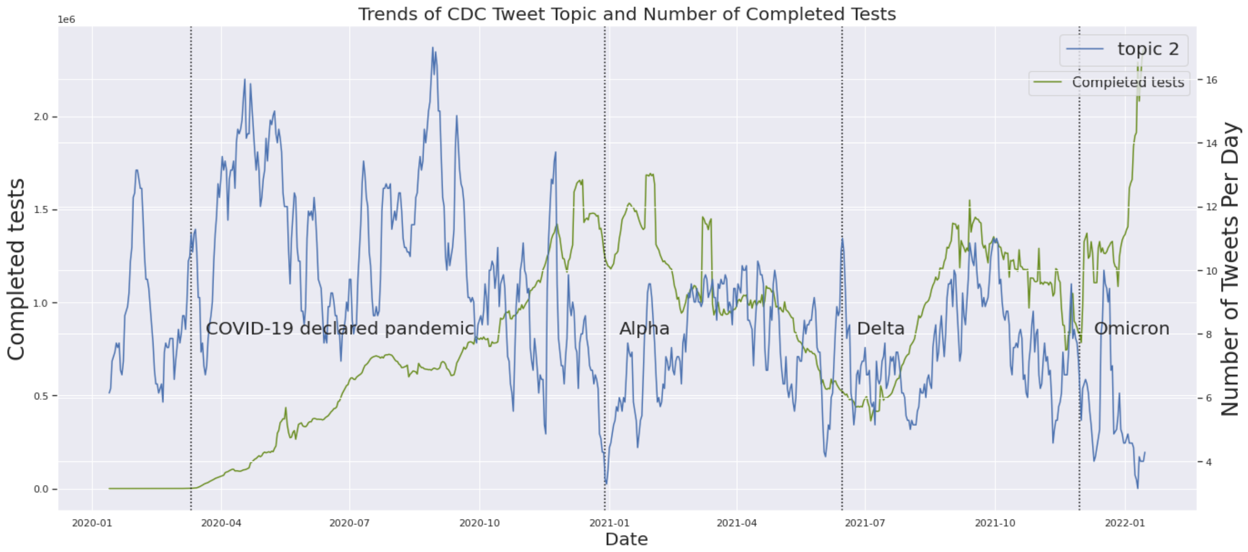


Figure 23.

- Tests and Topic 3:

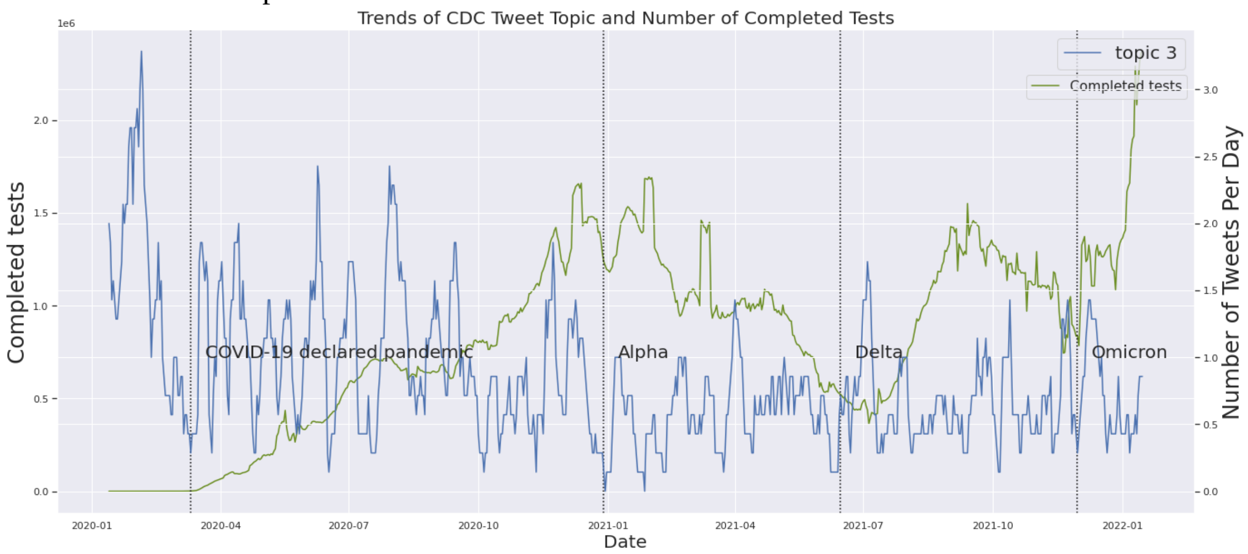


Figure 24.

- Tests and Topic 4:

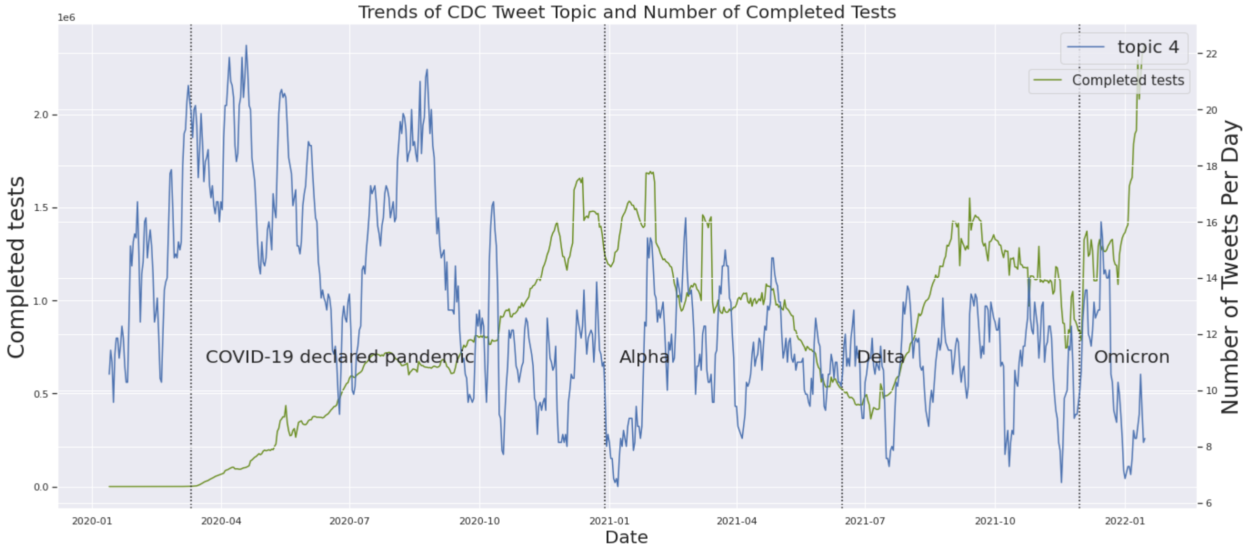


Figure 25.

- Vaccination and Topic 1:

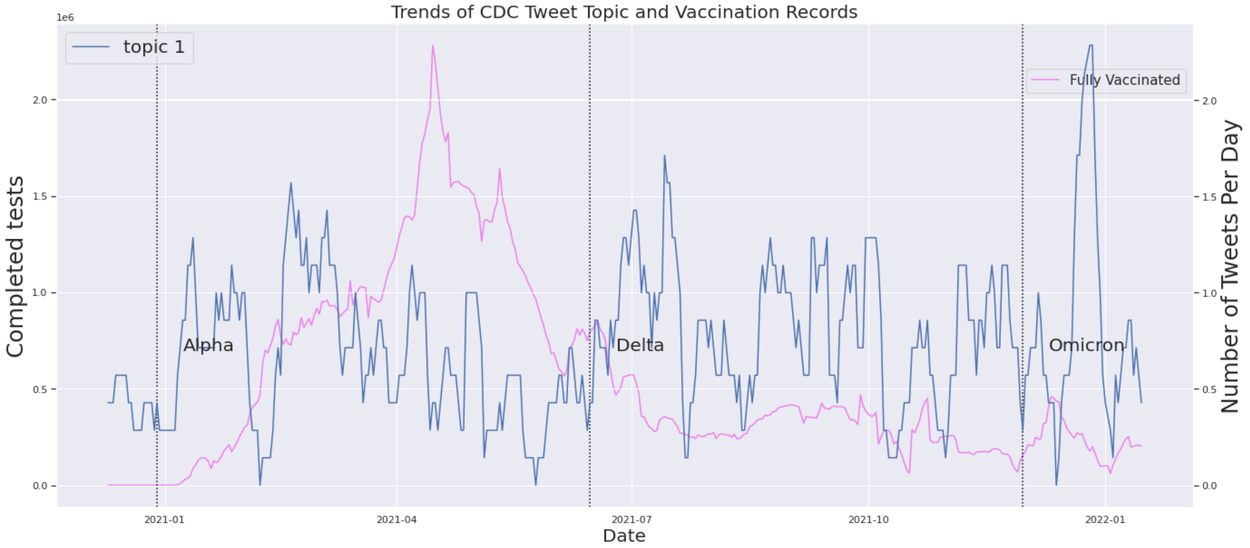


Figure 26.

- Vaccination and Topic 2:

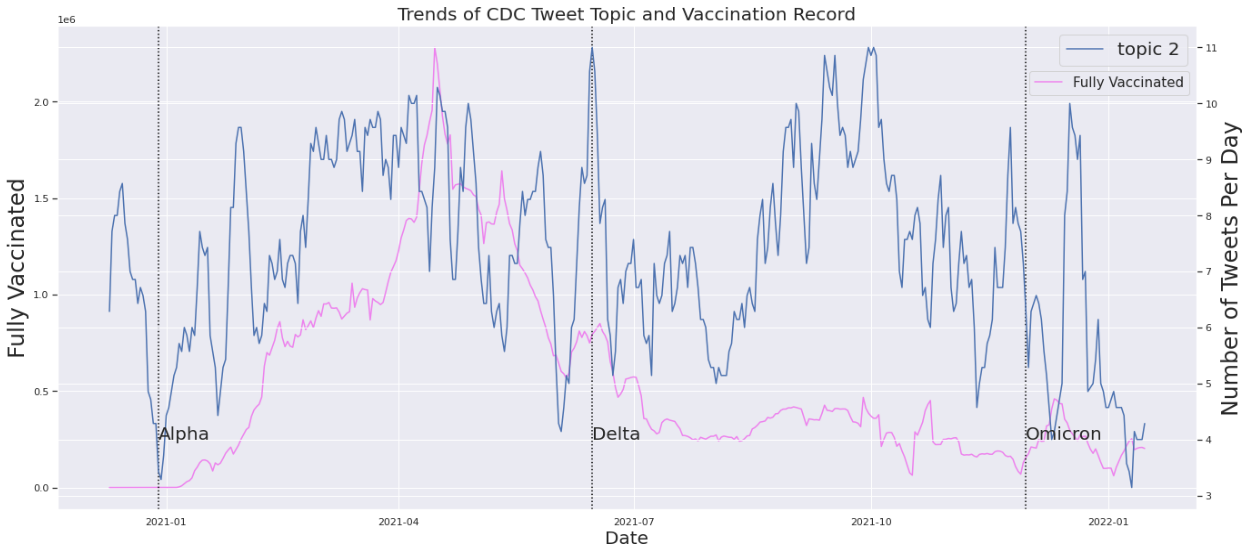


Figure 27.

- Vaccination and Topic 3

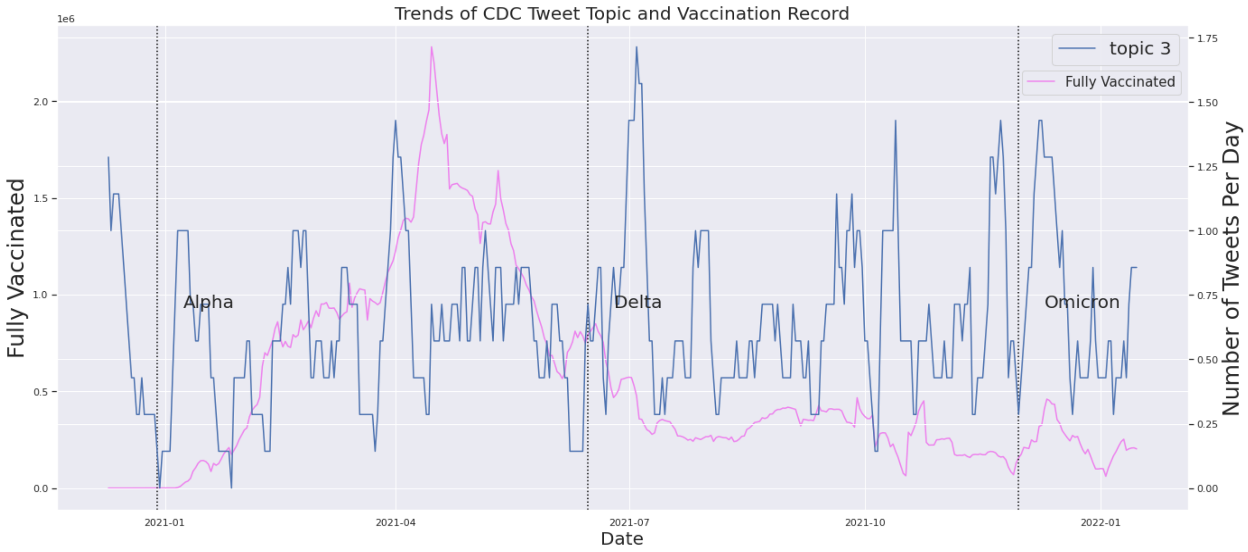


Figure 28.

- Vaccination and Topic 4:

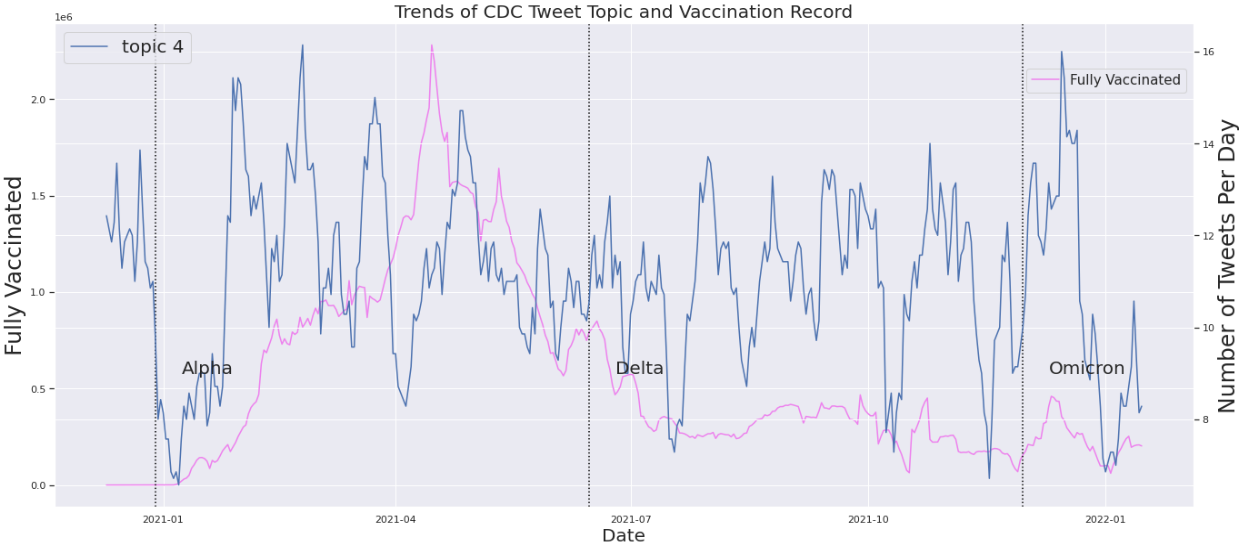


Figure 29.

**Cross correlation function (CCF) plots: CDC topics and COVID-19 epidemic measurements**

- Cases and topic 1

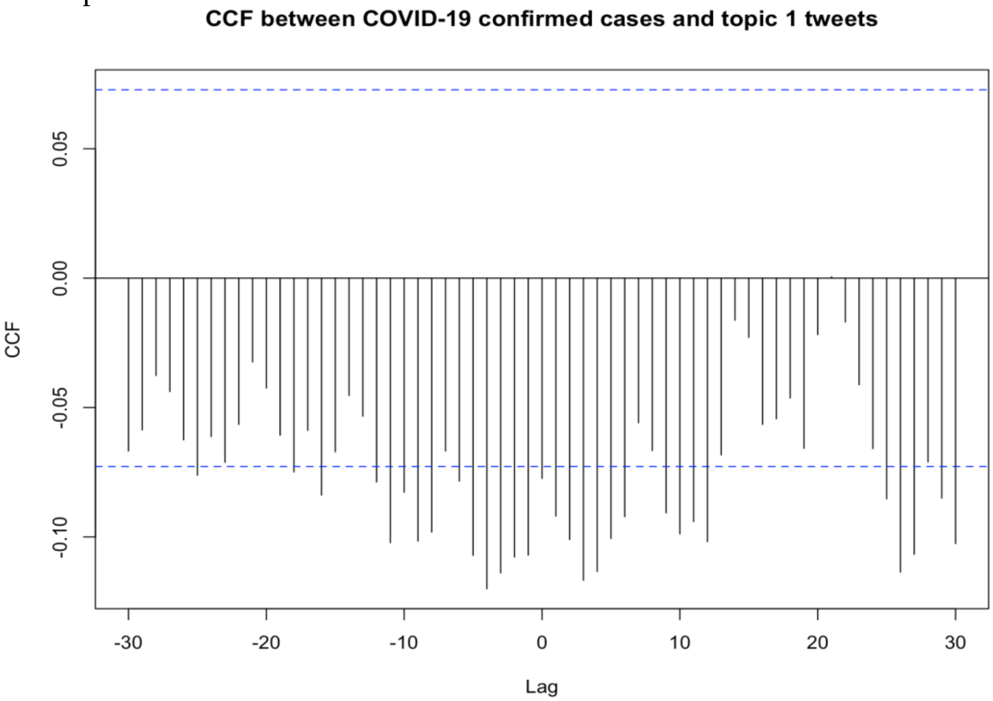


Figure 30.

- Cases and topic 2

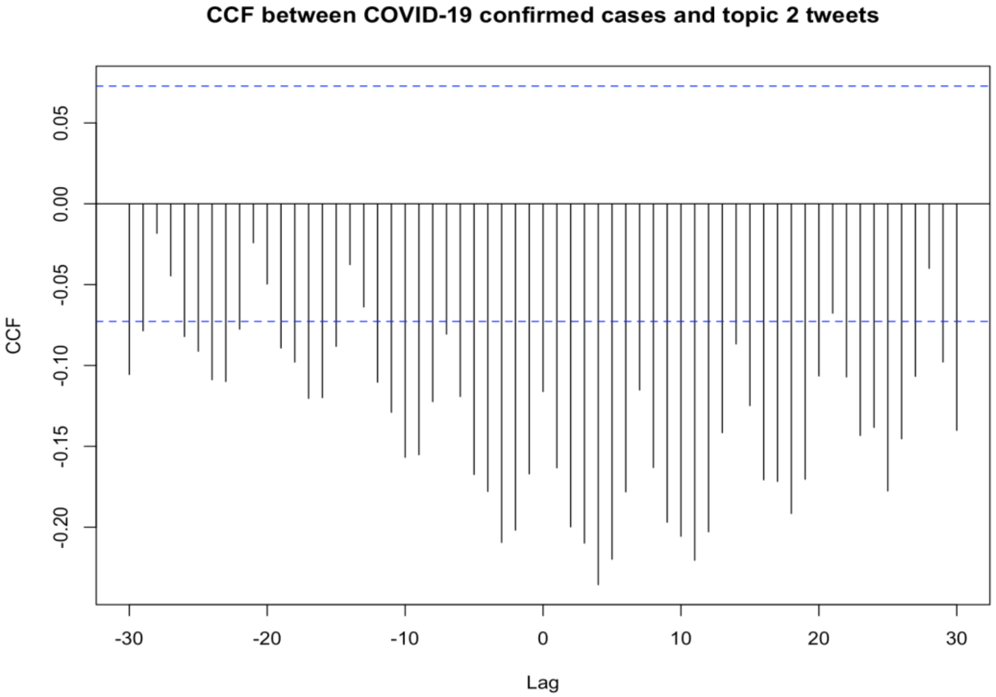


Figure 31.

- Cases and topic 3

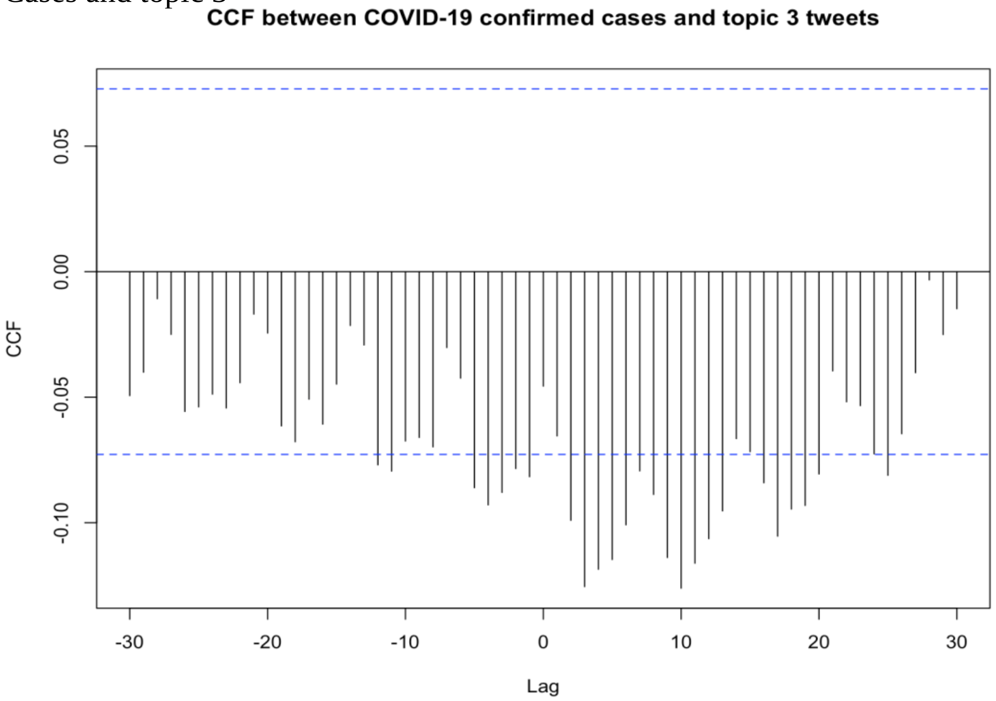


Figure 32.

- Cases and topic 4

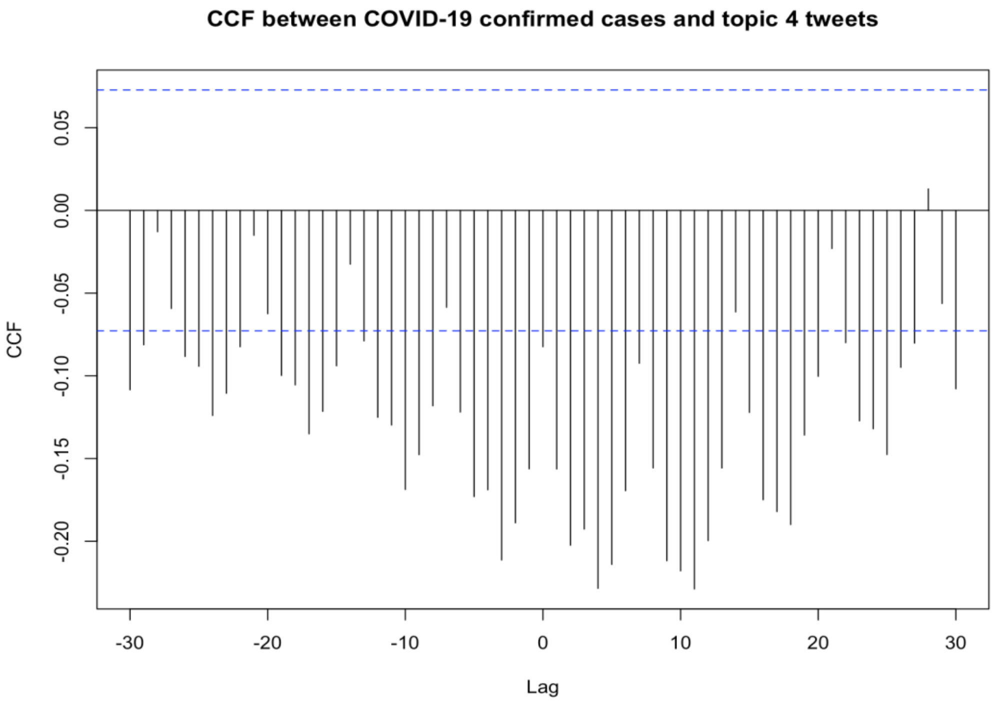


Figure 33.

- Deaths and topic 1

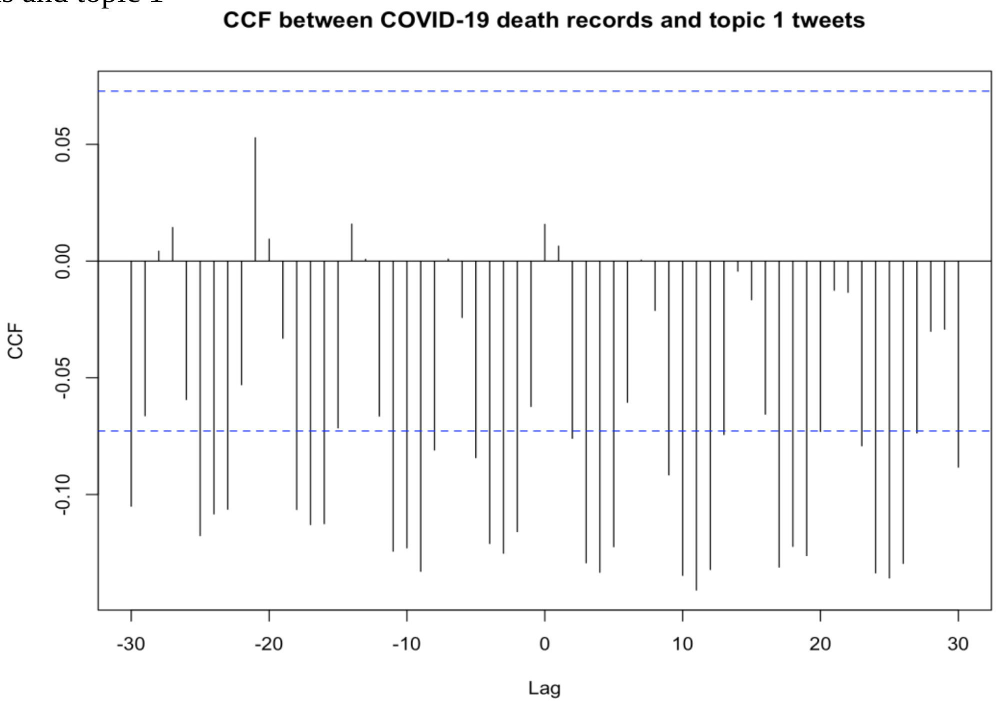


Figure 34.

- Deaths and topic 2

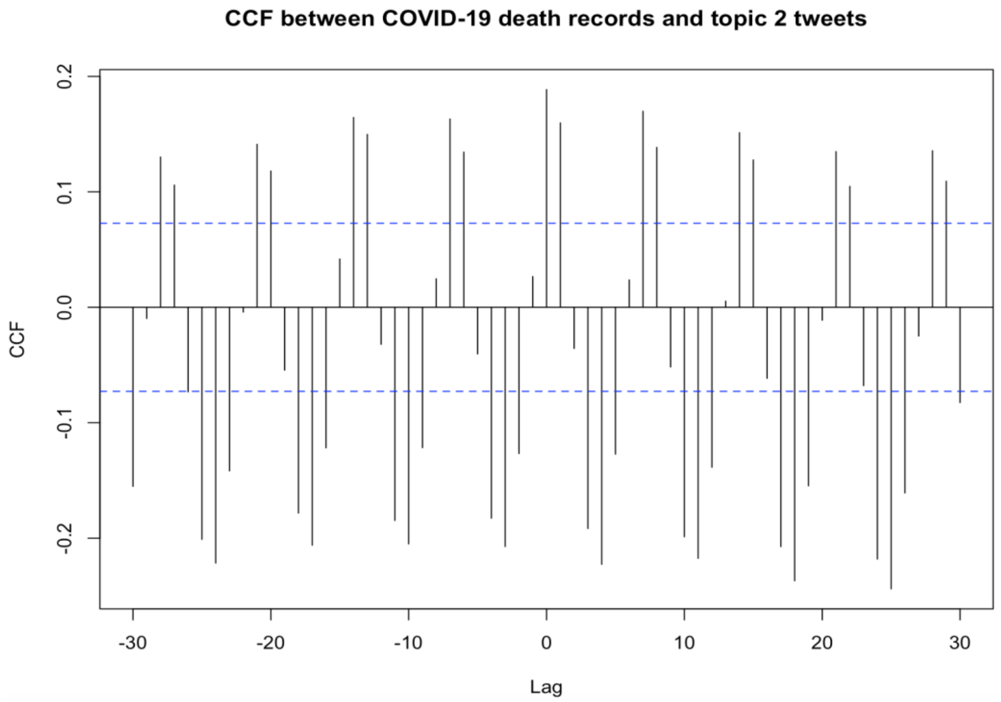


Figure 35.

- Deaths and topic 3

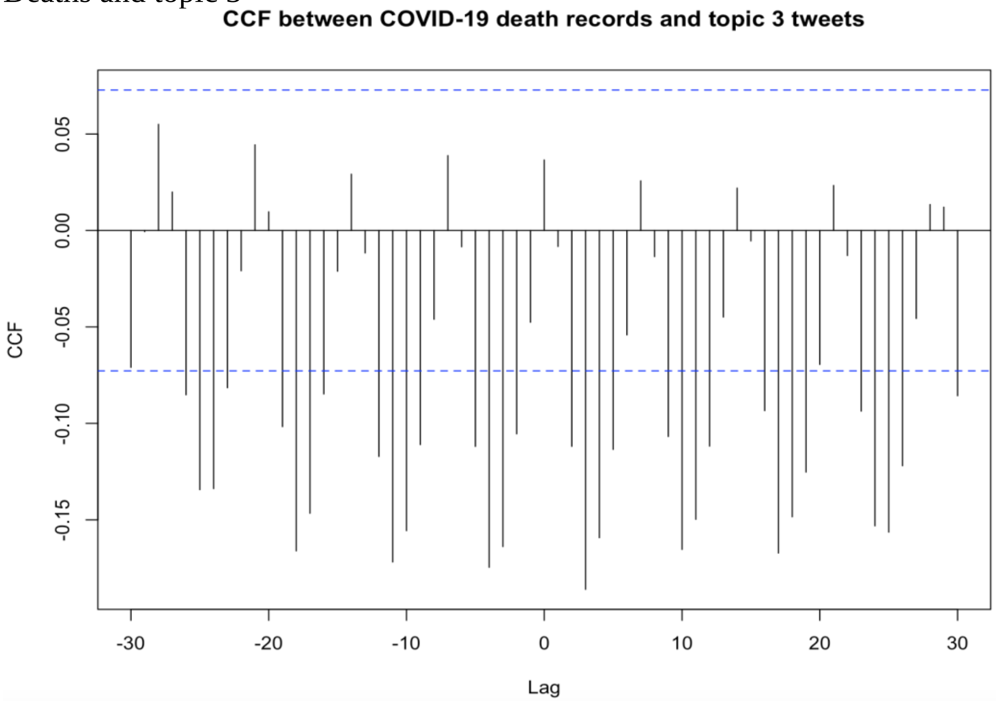


Figure 36.

- Deaths and topic 4

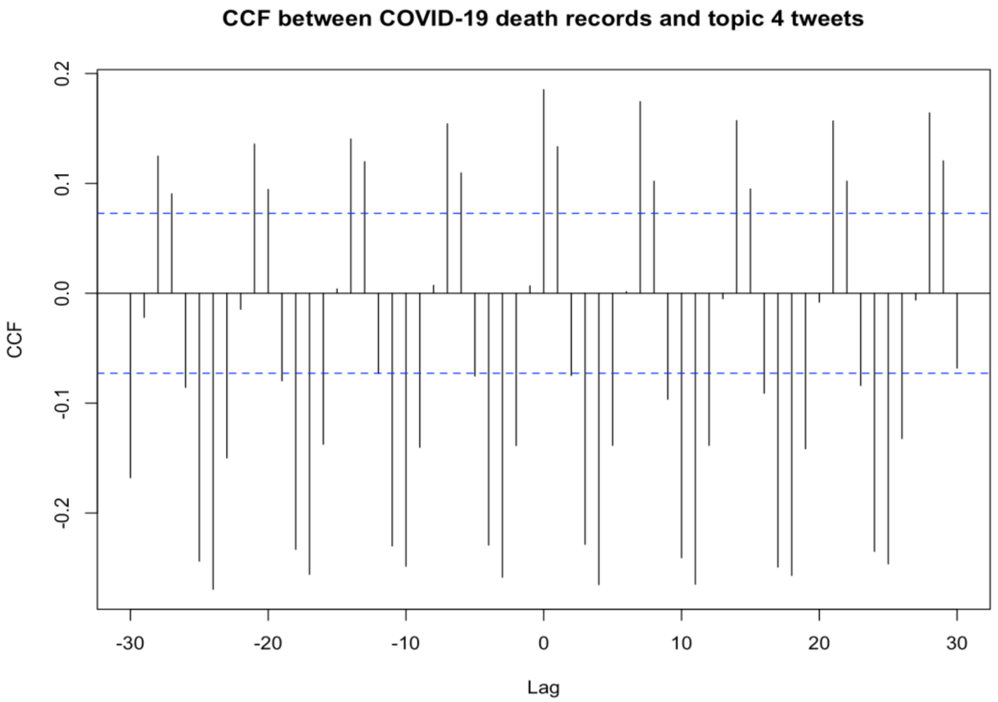


Figure 37.

- Testing and topic 1

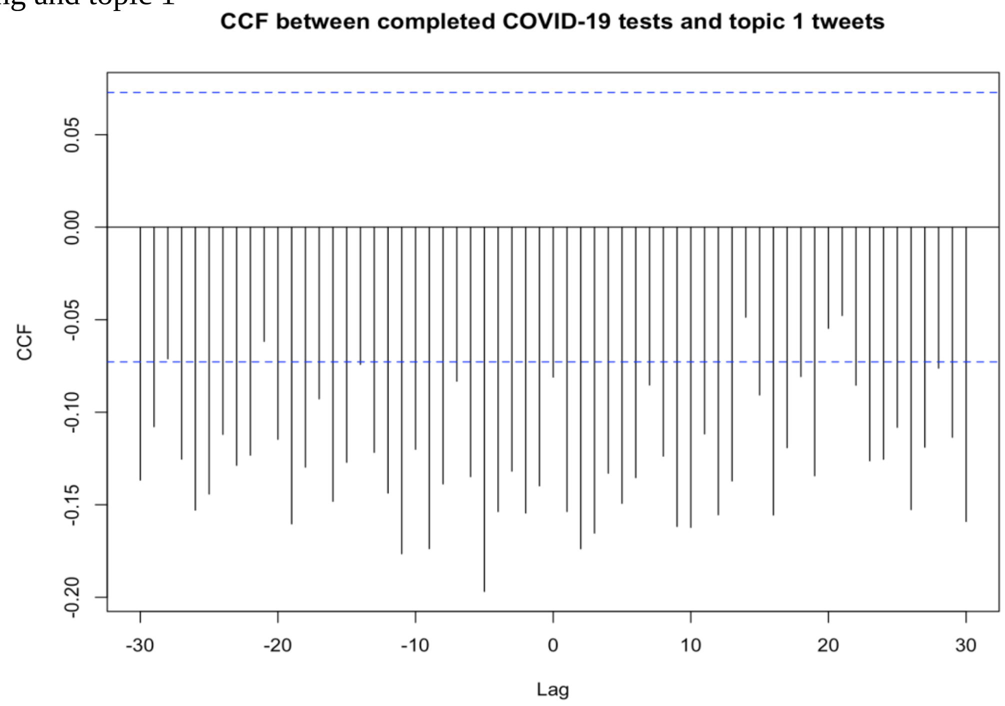


Figure 38.

- Testing and topic 2

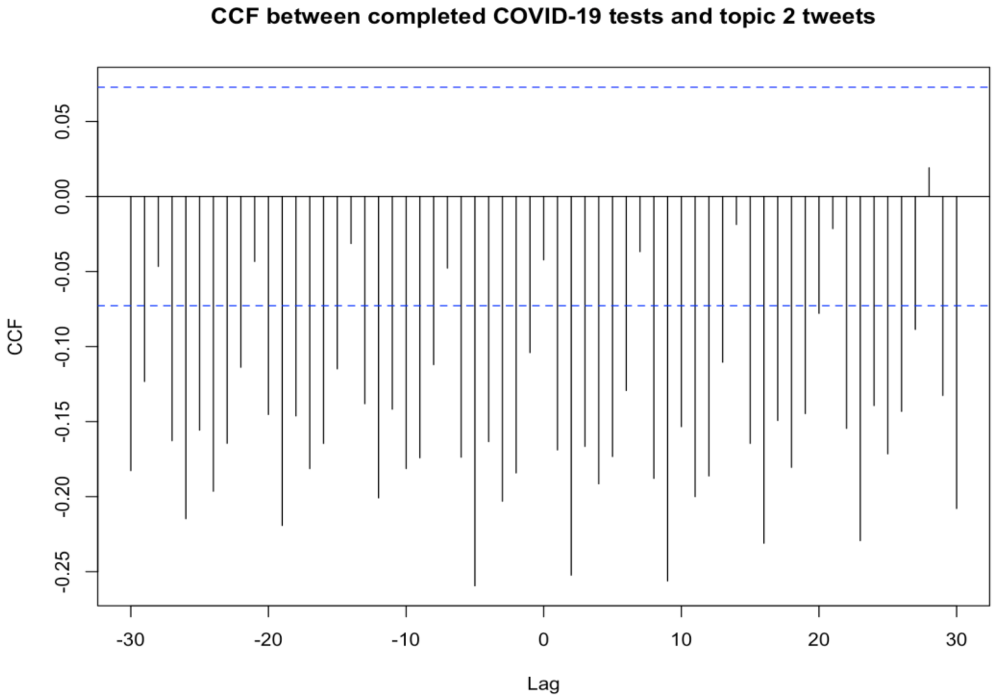

Figure 39.

- Testing and topic 3

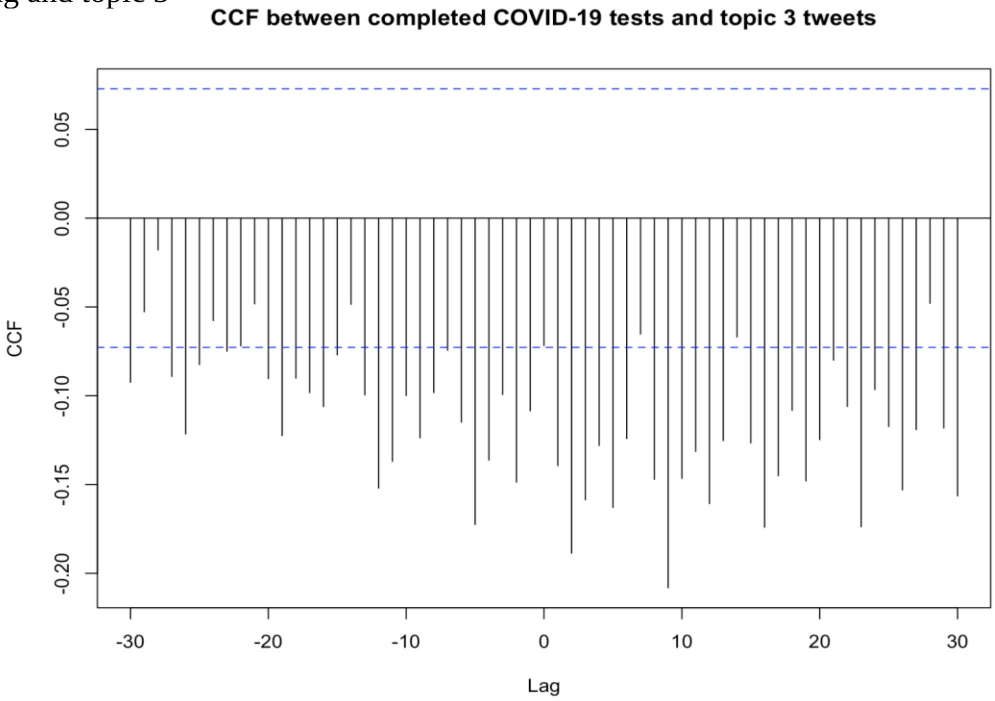

Figure 40.

- Vaccination and topic 1

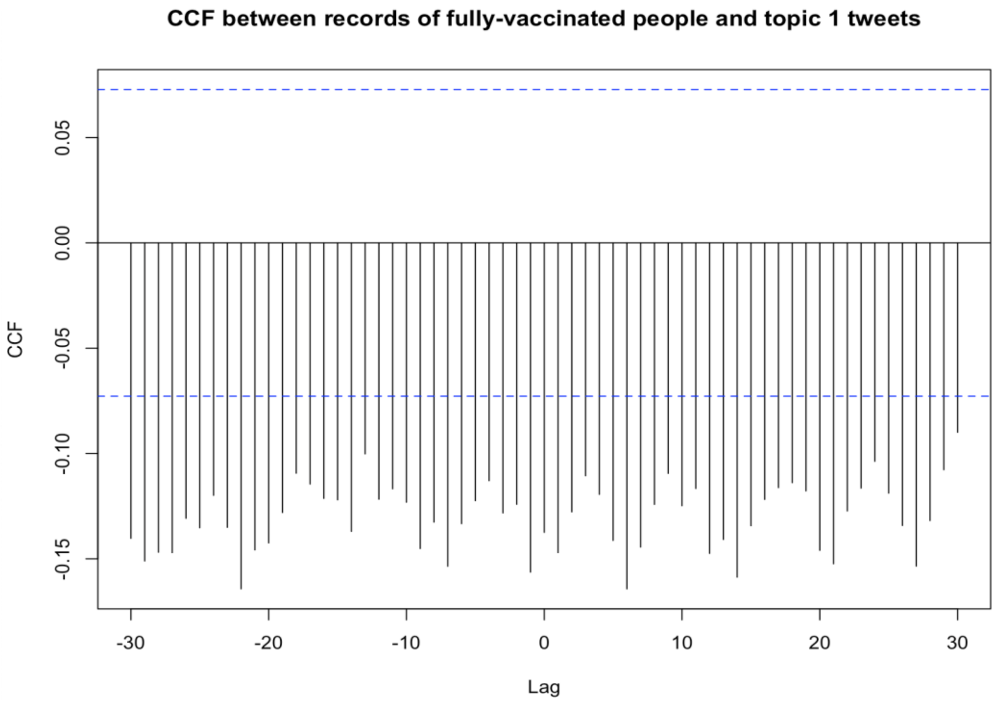


Figure 41.

- Vaccination and topic 2

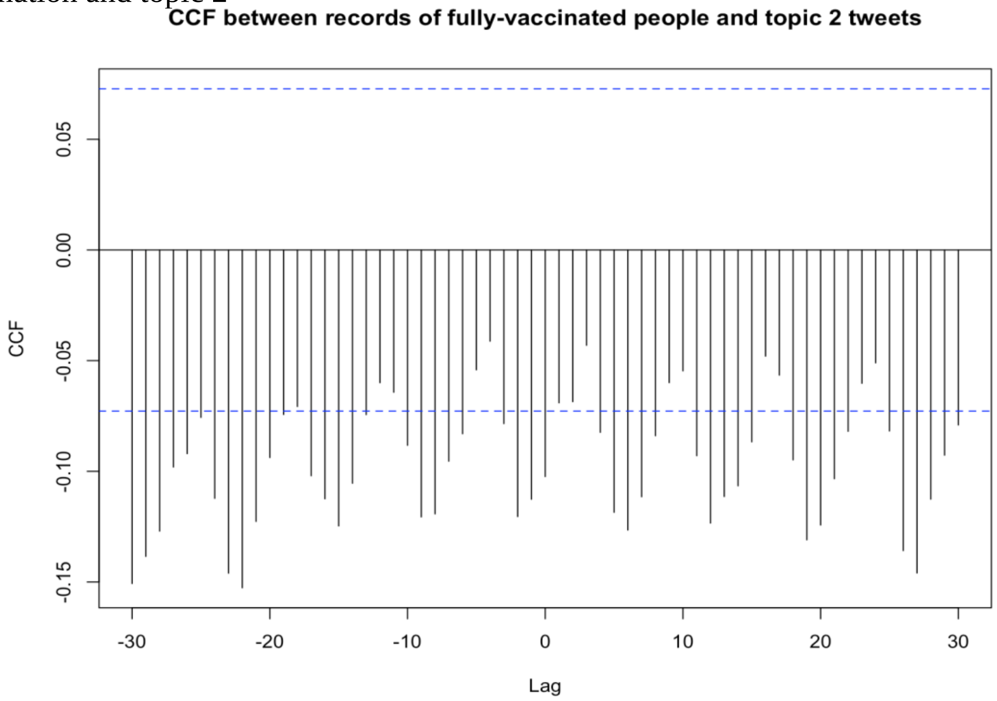


Figure 42.

- Vaccination and topic 3

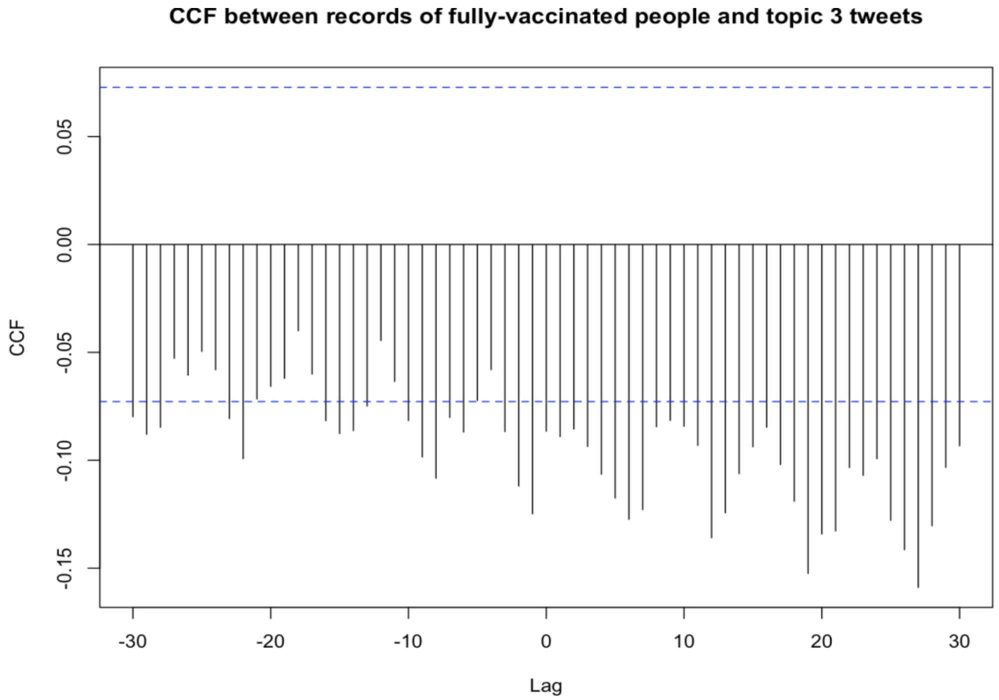


Figure 43.

- Vaccination and topic 4

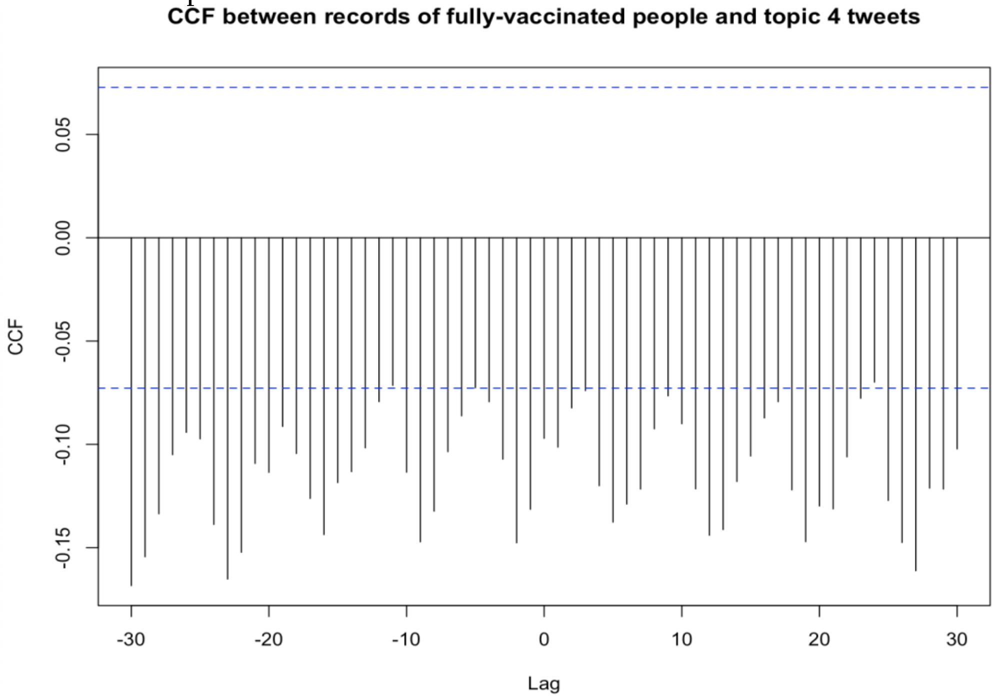


Figure 44.

**Maximum and minimum cross correlation functions (CCF) values and their lags between each CDC tweet topic series and each of the four COVID-19 epidemic metrics:**

**Table 3.** Cross correlations between CDC tweets from each topic and confirmed COVID-19 case counts in the United States from January 2020 to January 2022.

| Topic | Maximum CCF | Lag at Max | Minimum CCF | Lag at Min |
|---|---|---|---|---|
| 1 | 0.0005345247 | 21 | -0.1200439 | -4 |
| 2 | -0.01827029 | -28 | -0.2356 | 4 |
| 3 | -0.003315974 | 28 | -0.1260986 | 10 |
| 4 | 0.01299974 | 28 | -0.2288582 | 11 |

**Table 4.** Cross correlations between CDC tweets from each topic and COVID-19 death records in the United States from January 2020 to January 2022.

| Topic | Maximum CCF | Lag at Max | Minimum CCF | Lag at Min |
|---|---|---|---|---|
| 1 | 0.05283718 | -21 | -0.140962 | 11 |
| 2 | 0.1886017 | 31 | -0.2439371 | 25 |
| 3 | 0.05504533 | -28 | -0.1860584 | 3 |
| 4 | 0.1853307 | 0 | -0.2694168 | -24 |

**Table 5.** Cross correlations between CDC tweets from each topic and completed COVID-19 tests in the United States from January 2020 to January 2022.

| Topic | Maximum CCF | Lag at Max | Minimum CCF | Lag at Min |
|---|---|---|---|---|
| 1 | -0.04774661 | 21 | -0.1968656 | -5 |
| 2 | 0.01912121 | 28 | -0.2594286 | -5 |
| 3 | -0.01794747 | -28 | -0.2081833 | 9 |
| 4 | -0.008067961 | 28 | -0.3283732 | 9 |

**Table 6.** Cross correlations between CDC tweets from each topic and fully vaccinated records for COVID-19 in the United States from January 2020 to January 2022.

| Topic | Maximum CCF | Lag at Max | Minimum CCF | Lag at Min |
|---|---|---|---|---|
| 1 | -0.08994579 | 30 | -0.1642789 | 6 |
| 2 | -0.041227 | -4 | -0.1526447 | -22 |
| 3 | -0.03996313 | -18 | -0.1589435 | 27 |
| 4 | -0.06988632 | 24 | -0.1684011 | -30 |

**Mutual Information:**

**Table 7.** Mutual information between each topic in the United States from January 2020 to January 2022.

| Mutual Information | Topic 1 | Topic 2 | Topic 3 | Topic 4 |
|---|---|---|---|---|
| Topic 1 | 1.319797 | 0.1398849 | 0.06290801 | 0.1713954 |
| Topic 2 | 0.1398849 | 3.039821 | 0.1801499 | 0.7693809 |

| | | | | |
|---|---|---|---|---|
| Topic 3 | 0.06290801 | 0.1801499 | 1.255116 | 0.2175483 |
| Topic 4 | 0.1713954 | 0.7693809 | 0.2175483 | 3.338237 |
